# COVID-19 scenarios for the United States

**DOI:** 10.1101/2020.07.12.20151191

**Authors:** IHME COVID-19 Forecasting Team, Simon I Hay

## Abstract

The United States (US) has not been spared in the ongoing pandemic of novel coronavirus disease^1,2^. COVID-19, caused by the severe acute respiratory syndrome coronavirus 2 (SARS-CoV-2), continues to cause death and disease in all 50 states, as well as significant economic damage wrought by the non-pharmaceutical interventions (NPI) adopted in attempts to control transmission^3^. We use a deterministic, Susceptible, Exposed, Infectious, Recovered (SEIR) compartmental framework^4,5^ to model possible trajectories of SARS-CoV-2 infections and the impact of NPI^6^ at the state level. Model performance was tested against reported deaths from 01 February to 04 July 2020. Using this SEIR model and projections of critical driving covariates (pneumonia seasonality, mobility, testing rates, and mask use *per capita*), we assessed some possible futures of the COVID-19 pandemic from 05 July through 31 December 2020. We explored future scenarios that included feasible assumptions about NPIs including social distancing mandates (SDMs) and levels of mask use. The range of infection, death, and hospital demand outcomes revealed by these scenarios show that action taken during the summer of 2020 will have profound public health impacts through to the year end. Encouragingly, we find that an emphasis on universal mask use may be sufficient to ameliorate the worst effects of epidemic resurgences in many states. Masks may save as many as 102,795 (55,898–183,374) lives, when compared to a plausible reference scenario in December. In addition, widespread mask use may markedly reduce the need for more socially and economically deleterious SDMs.

---

The zoonotic origin of the novel severe acute respiratory syndrome coronavirus 2 (SARS-CoV-2)^7^ in Wuhan, China^8^, and the global spread of the coronavirus disease (COVID-19)^2,9^ promises to be the defining global health event of the twenty-first century. This pandemic has already resulted in extreme societal, economic, and political disruption across the world and in the United States (US)^3,10^. The establishment of SARS-CoV-2 and its rapid spread in the US has been dramatic^11^. Since the first case in the US was identified on 20 January 2020^12^ (first death on 06 February 2020^13^), SARS-CoV-2 has spread to every state and resulted in more than 15.7 million cases and 127,868 deaths as of 4 July 2020^14–16^.

There remains no approved vaccine for the prevention of SARS-CoV-2 infection and few pharmaceutical options for the treatment of the COVID-19 disease^17,18^. The most optimistic commentators do not predict the availability of new vaccines or therapeutics before 2021^19^. Non-pharmaceutical interventions (NPI) are, therefore, the only available policy levers to reduce transmission^20^. Several such NPI have been put in place across the US in response to the epidemic (Fig. 1), including the dampening of transmission through the wearing of face masks and social distancing mandates (SDM) aimed at reducing contacts through school closures, restrictions of gatherings, stay at home orders, and the partial or full closure of non-essential businesses. Increased testing and isolation of infected individuals will also have had an impact^6^. These NPI are credited with a reduction in disease transmission^21,22^, along with a host of other hypotheses on environmental, behavioral, and social determinants of the course of the epidemic at the state level.

**Fig. 1.**
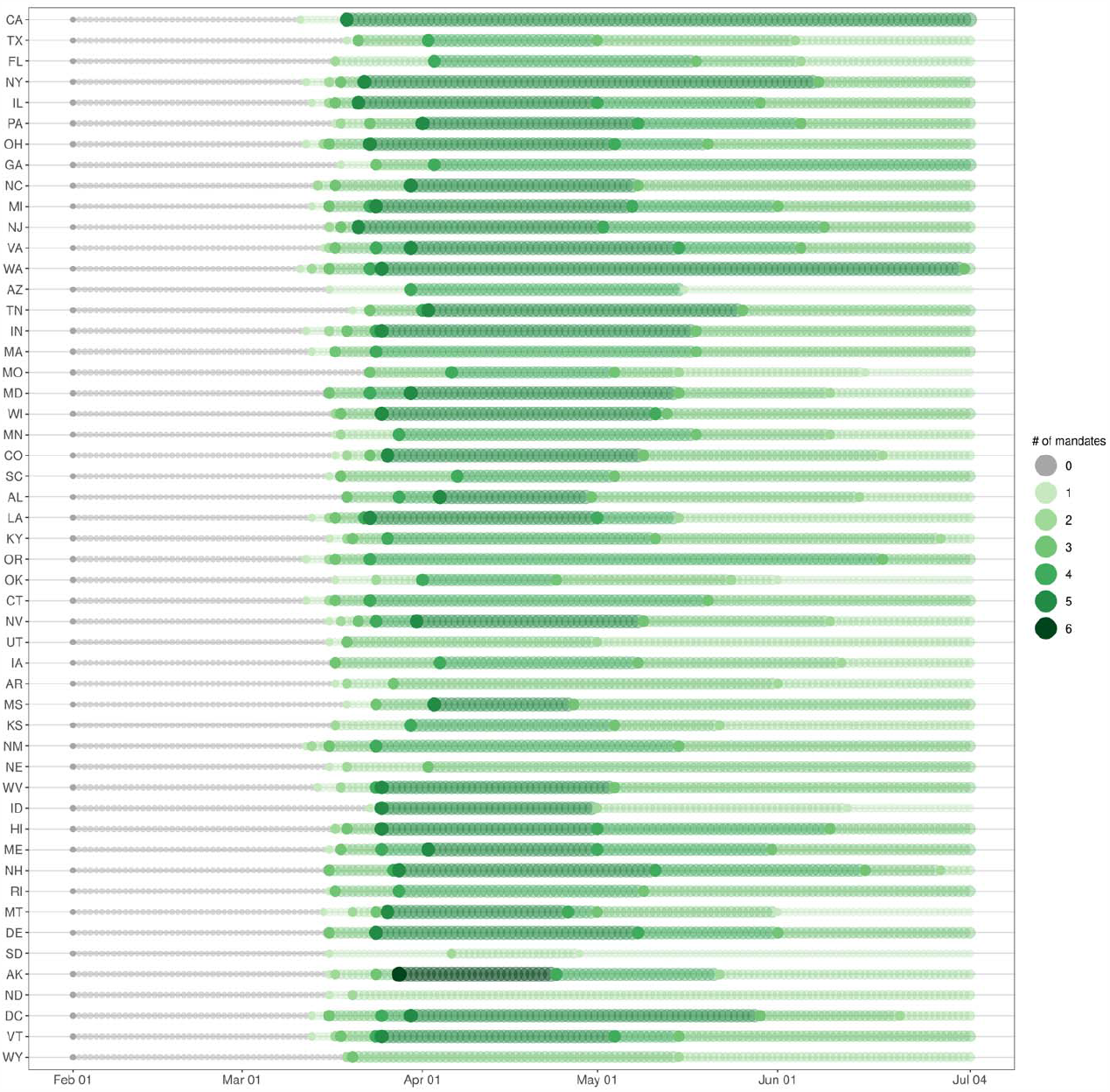
Number of social distancing mandates by state in the US on a timeline starting on 01 February 2020 through to July 04 2020. States are ordered by decreasing population size on the y-axis.

In the US, decisions to impose SDM or require mask use are generally made at the state level by government officials. These executives need to balance net losses from the societal turmoil, economic damage, and indirect effects on health caused by NPI with the direct benefits to human health of controlling the epidemic, all within a complex political environment. Control has usually been defined as the restriction of infections to below a specified level at which health services are not overwhelmed by demand and the loss of human health and life is minimized^23^.

In the first stages of the SARS-CoV-2 outbreak in the US, states sequentially enacted increasingly restrictive SDMs meant to reduce transmission (by reducing human-to-human contact)^1^ at the same time as there was conflicting advice on the use of masks by the general public^24^. At that early stage, relatively simple statistical models of future risk were sufficient to capture the general patterns of transmission^25^. As different behavioral responses to SDM began to emerge, and more importantly, as some states began to remove SDM (Fig. 1), a modeling approach that directly quantifies transmission and could be used to explore these developing scenarios was necessary^25^. As states variously remove and reinstate SDM (Fig. 1) or begin to issue mandatory mask use orders^26^ amid resurgences of COVID-19^27^, there is an urgent need for evidence-based assessments of the likely impact of the NPI options available to decision-makers.

There is now a growing consensus that face masks, whether cloth or medical-grade, can considerably reduce the transmission of respiratory viruses like SARS-CoV2, thereby limiting spread of COVID-19^28–30^. While medical-grade masks may provide enhanced protection, cloth face coverings (homemade or manufactured), have been found to be comparably effective in non-medical settings^28^, as well as being simple, widely accessible, and available commonly at relatively low cost. We updated a recently published review^28^ to generate a novel meta-analysis (Supplementary Information section 3.4) of both peer-reviewed studies and pre-prints to assess mask effectiveness at preventing respiratory viral infections in humans^31^. This analysis suggested a reduction in infection (from all respiratory viruses) for mask-wearers by one-third (Relative Risk = 0.65 (0.47-0.92)) relative to controls. This is suggestive of a considerable population health benefit to mask wearing that may be particularly effective in the US, where currently only 41.1% of Americans have reported always wearing a mask in public (Supplementary Information section 3.4)^32^.

Here we provide a state-level descriptive epidemiological analysis of the introduction of SARS-CoV-2 infection across the US, from the first recorded case, through to 04 July 2020. We use these observations to learn about epidemic progression and thereby model the first wave of transmission using a deterministic Susceptible, Exposed, Infectious, Recovered (SEIR) compartmental framework^4,5^. This observed, process-based understanding of how NPI affect epidemiological processes is then used to make inferences about the future trajectory of COVID-19 and how different combinations of existing NPI might affect this course. Three SEIR-driven scenarios, along with covariates that inform them, were then projected until 31 December 2020 (see methods). We use these scenarios as a sequence of experiments to describe a range of model outputs including *R*_*effective*_ (the change over time in the average number of secondary cases per infectious case in a population where not everyone is susceptible^4,5,33^), infections, deaths, and hospital demand outcomes which might be expected from plausible subsets of the policy options applied in the summer and fall of 2020 (see methods, Supplementary Information section 6.1 for more rationale on scenario construction and considerations).

Briefly, we forecast the expected outcomes if states continue to remove SDMs at the current pace (“mandates easing”), with resulting increases in population mobility and number of contacts. This is an alternative scenario to the more probable situation, where states are expected to respond to an impending health crisis by re-imposing some SDMs. In that plausible reference scenario, we model the future progress of the pandemic assuming that states would move to once again shut down social interaction and economic activity at a threshold for the daily death rate; when 8 daily deaths per million population is reached – the 90^th^ percentile of the observed distribution of when states previously implemented SDM (Fig. 1, Supplementary Information section 3) – we assume reinstatement of SDM for six weeks. In addition, newly available data on mask efficacy enabled the exploration of a third, “universal mask” scenario to investigate the potential population-level benefits of increased mask use in addition to a threshold-driven reinstatement of SDM. In this scenario, “universal” was defined as 95% of people wearing masks in public, based on the current highest rate of mask use globally (in Singapore), during the COVID-19 pandemic to date (Supplementary Information section 3.4). All scenarios presume an increase in mobility associated with the opening of schools across the country.

## Observed COVID-19 trends

The COVID-19 epidemic has progressed unevenly across states. Since the first death was recorded in the US in early February 2020, cumulative through 04 July 2020, 127,868 deaths from COVID-19 have been reported in the US (Fig. 2); a quarter of those (24.5%) occurred in New York alone. Washington and California issued the first sets of state-level mandates on 11 March that prohibited gatherings of 250 people or more in certain counties, and by 23 March, all 50 states initiated some combination of SDM (Fig. 1). The highest levels of daily deaths at the state level between February and June of 2020 occurred in New York, New Jersey, and Massachusetts at 935.3, 330.2, and 168.1 deaths per day (Fig. 3, Extended Data Fig. 1). At the end of June, the highest level of daily deaths was in California at 73.5 deaths per day. A critical policy need at this stage of the modeling was the forecasting of hospital demand in the US in the states with the worst effective transmission rates (Hawaii, South Carolina, and Florida; Fig. 4). The highest peak demand was observed as 5969 hospital ICU beds in New York on April 8 and 3073 ICU beds in New Jersey on April 19; health care capacity was exceeded in 11 states (New York, New Jersey, Connecticut, Massachusetts, Michigan, Maryland, Louisiana, Pennsylvania, Rhode Island, Delaware, District of Columbia) (Extended Data Figs 2,3). Demand had receded to within capacity levels across the US by the end of May.

**Fig. 2.**
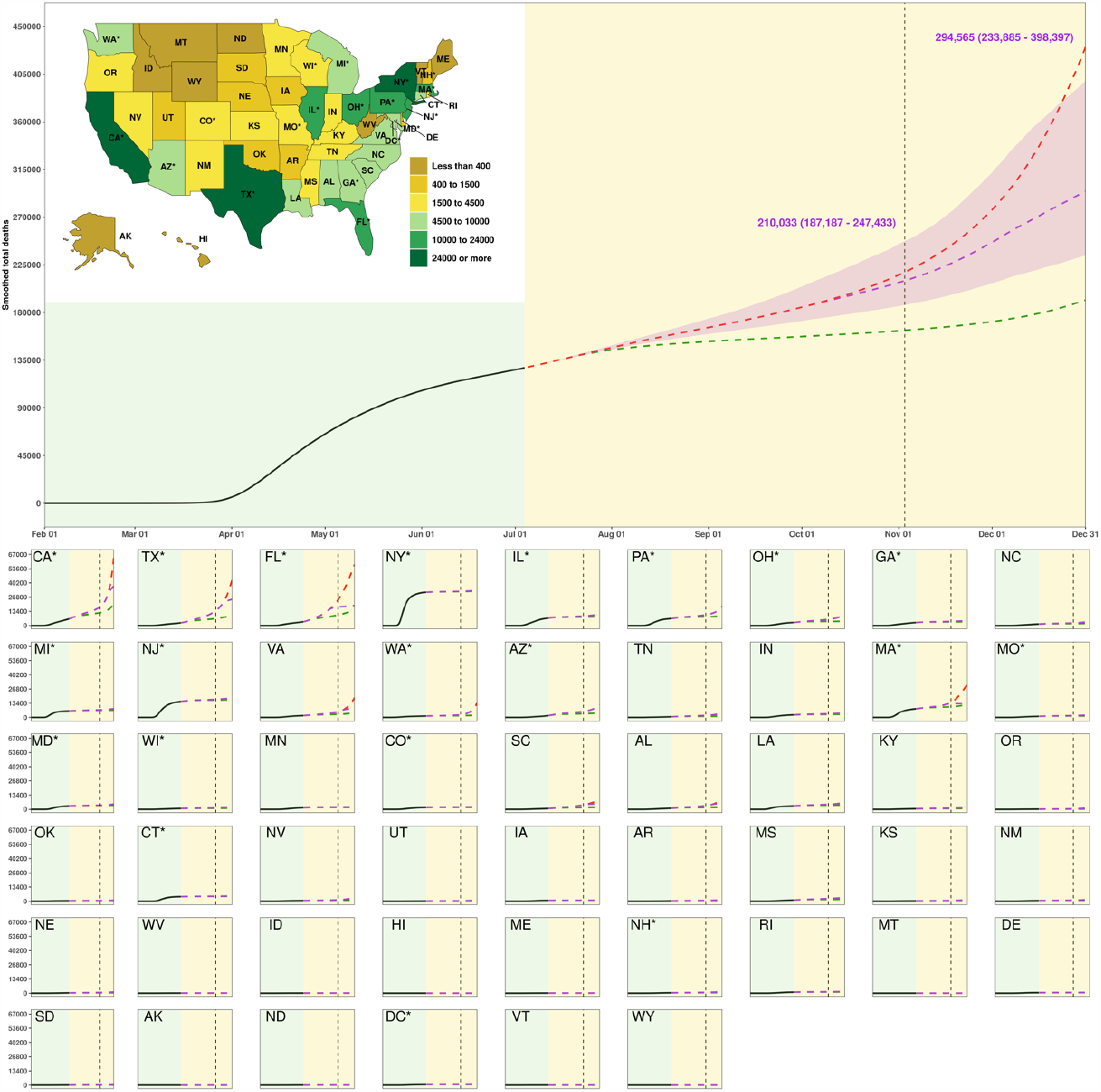
Cumulative deaths from 01 February to 31 December 2020. The inset map displays the cumulative deaths under the “plausible reference” scenario on 31 December 2020. A light yellow background separates the observed and predicted part of the time series, before and after 04 July. The dashed vertical line is 03 November. The red line is the “mandates easing” scenario, the purple line the “plausible reference” scenario, and green line the “universal mask” scenario. Numbers are the means and UIs for the plausible reference scenario on dates highlighted. The UIs are not shown for “mandates easing” and mask use scenario for clarity. State panels are ordered by decreasing population size. Two-letter state abbreviations are provided in panels and the inset map. An asterisk next to state abbreviation indicates a state with one or more urban agglomerations exceeding two million persons. State panels are scaled to accommodate the state with the highest value (CA here), ranging from zero to 68,000 cumulative deaths. This map was generated with RStudio (R Version 3.6.3).

**Fig. 3.**
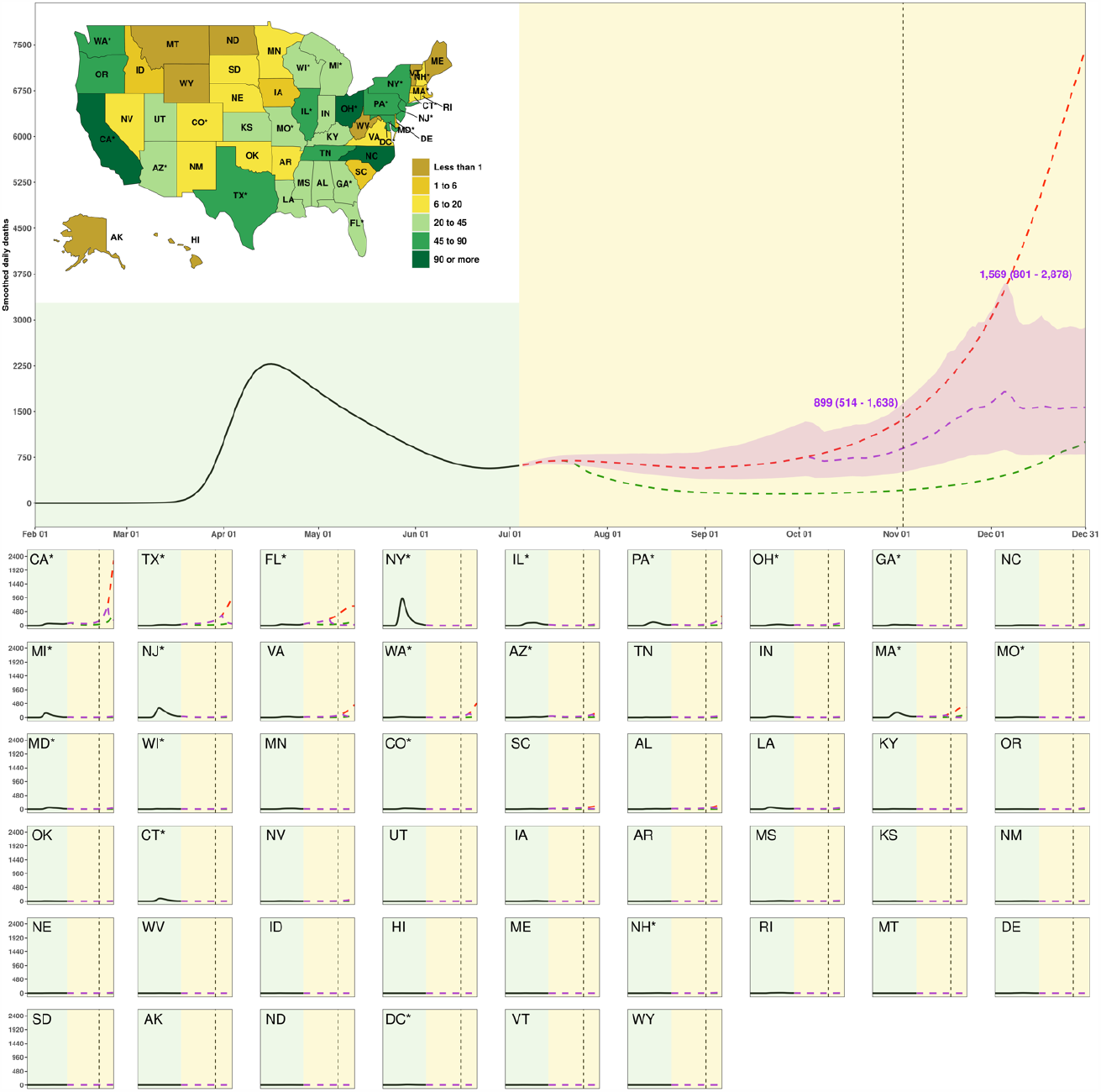
Daily deaths from 01 February to 31 December 2020. The inset map displays the daily deaths under the “plausible reference” scenario on 31 December 2020. A light yellow background separates the observed and predicted part of the time series, before and after 04 July. The dashed vertical line is 03 November. The red line is the “mandates easing” scenario, the purple line the “plausible reference” scenario, and the green line the “universal mask” scenario. Numbers are the means and UIs for the plausible reference scenario on dates highlighted. The UIs are not shown for the “mandates easing” and “universal mask” scenarios for clarity. State panels are ordered by decreasing population size. Two-letter state abbreviations are provided in panels and the inset map. An asterisk next to state abbreviation indicates a state with one or more urban agglomerations exceeding two million persons. State panels are scaled to accommodate the state with the highest value (CA here), ranging from zero to 2,500 daily deaths. This map was generated with RStudio (R Version 3.6.3).

**Fig. 4.**
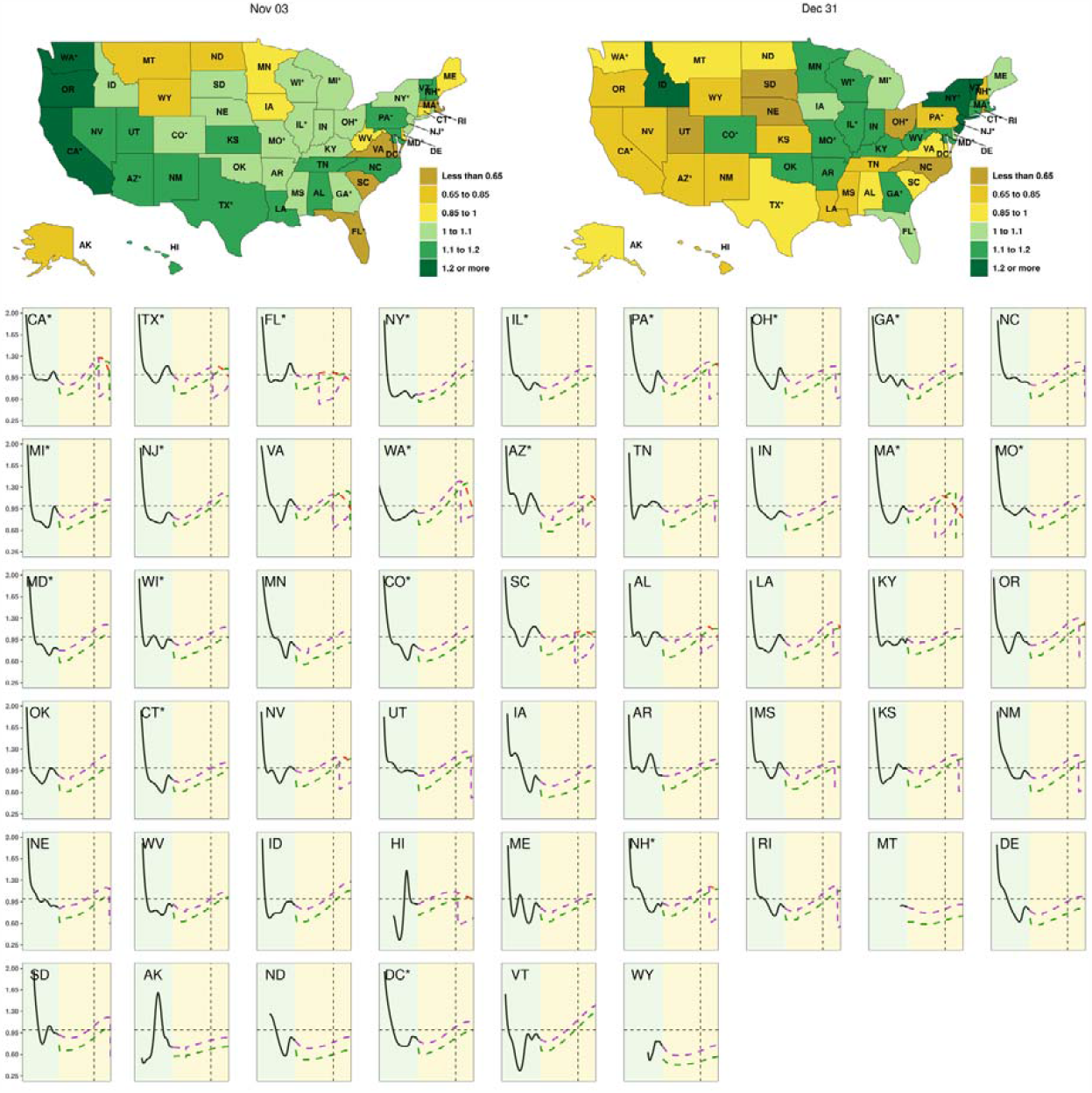
Time series for values of R_effective_ by state in the US. Inset maps display the value of R_effective_ on 03 November and 31 December 2020; time series of R_effective_ are presented for each state as separate panels. Time series for values of R_effective_ by state in the US. Inset maps display the value of R_effective_ on 03 November and 31 December 2020; time series of R_effective_ are presented for each state as separate panels. A light yellow background separates the observed and predicted part of the time series, before and after 04 July. The dashed vertical line is 03 November. The red line is the “mandates easing” scenario, the purple line the “plausible reference” scenario, and green line the “universal mask” scenario. The UIs are not shown for “mandates easing” and mask use scenario for clarity. State panels are ordered by decreasing population size. Two-letter state abbreviations are provided in panels and the inset maps. An asterisk next to state abbreviation indicates a state with one or more urban agglomerations exceeding two million persons. For legibility purposes, the y-axes of the state panels go from 0.25 to 2 and the x-axes go from 01 March to 31 December. These maps were generated with RStudio (R Version 3.6.3).

## Predicted COVID-19 trends

Under a scenario where states continue with planned removal of SDMs (“mandates easing”), our model projects that cumulative total deaths across the US could reach 430,494 (288,046–649,582) by 31 December 2020 (Fig. 2, Table 1). At the state level, contributions to that death toll would not be evenly distributed across the US. Greater than 60% of the deaths projected between July and December 2020 in this scenario would occur across just five states: California, Florida, Texas, Massachusetts, and Virginia; the highest cumulative death rates (per 100,000) between July and December 2020 are projected to occur in Massachusetts (465.0 (302.4–659.9) deaths per 100,000)), Florida (272.4 (117.3– 551.0) deaths per 100,000), Virginia (214.9 (78.4–468.8) deaths per 100,000), and New Jersey (207.2 (191.5-235.0) deaths per 100,000) (Extended Data Fig. 4, Table 1). By 03 November 2020 – when many Americans may need to queue in public for national elections – a total of four states are predicted to exceed a threshold of daily deaths of 8 deaths per million (Fig. 3), and a total of 41 states would have an *R*_*effective*_ greater than one (Fig. 4), presenting a possible increased risk of spread if preventive measures are not taken at that time. By 31 December 2020, a total of 24 states are predicted to exceed that threshold and 47 states would reach an *R*_*effective*_ of greater than one before the end of the year (Table 1; Fig. 4). This scenario results in an estimated total of 67,485,279 (41,003,799–101,794,827) infections across the United States by the end of year (Extended Data Fig. 5). The highest infection levels in states relative to their population are estimated to occur in Massachusetts (58.0% (39.9–74.9%) infected), Virginia (37.5% (13.8–68.0%) infected), and Washington (37.1% (15.0–67.0%) infected) (Extended Data Fig. 6). Further results for hospital resource use needs are presented in Extended Data Figs 2,3 and forecast infections under this scenario are presented in Extended Data Figs 7,8.

**Table 1.**
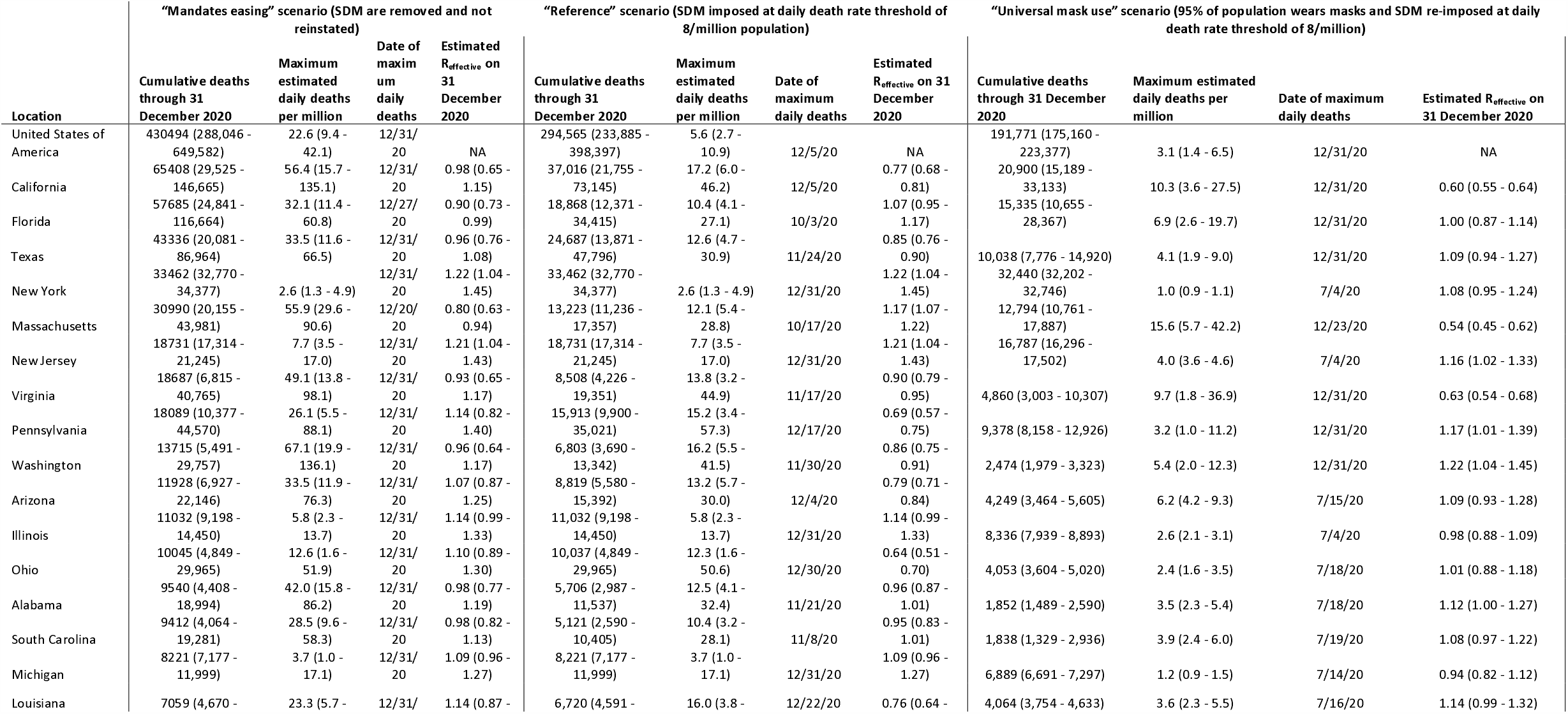
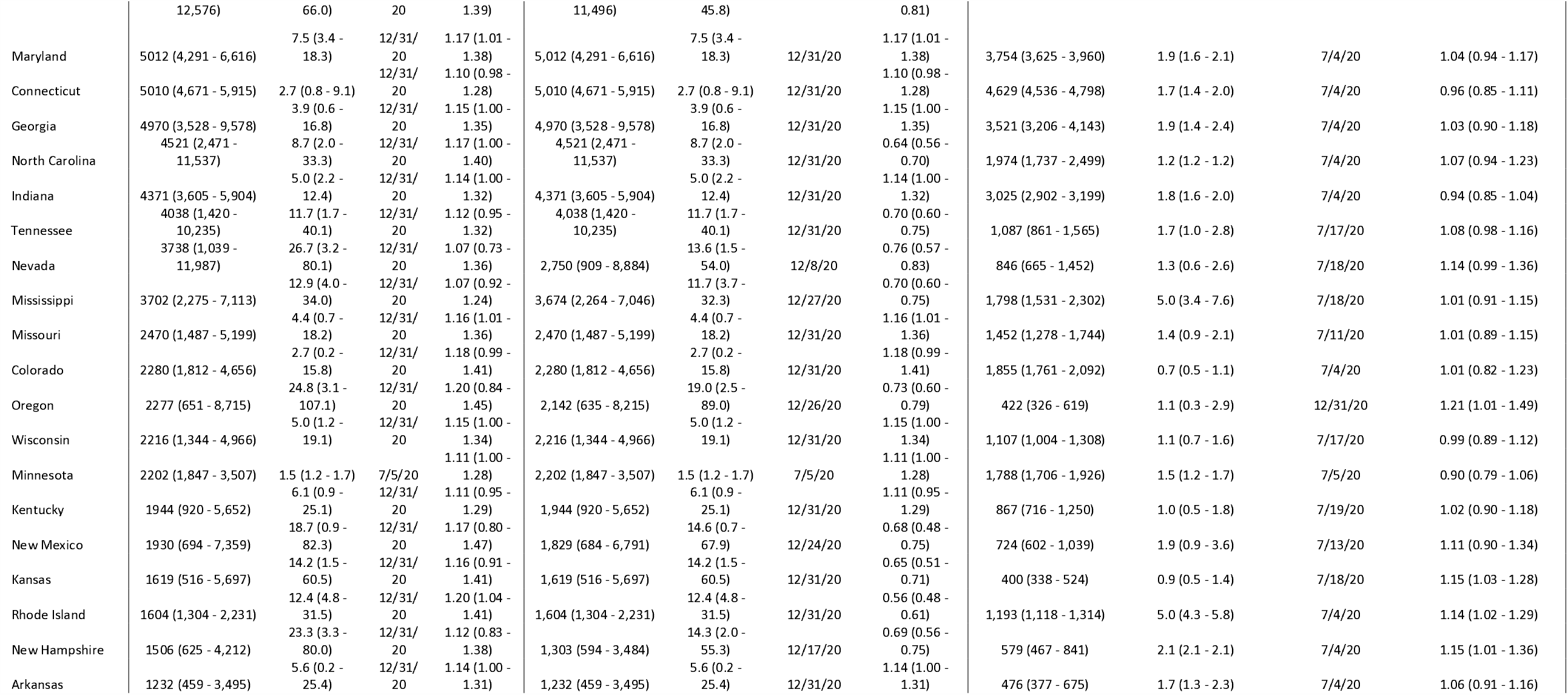
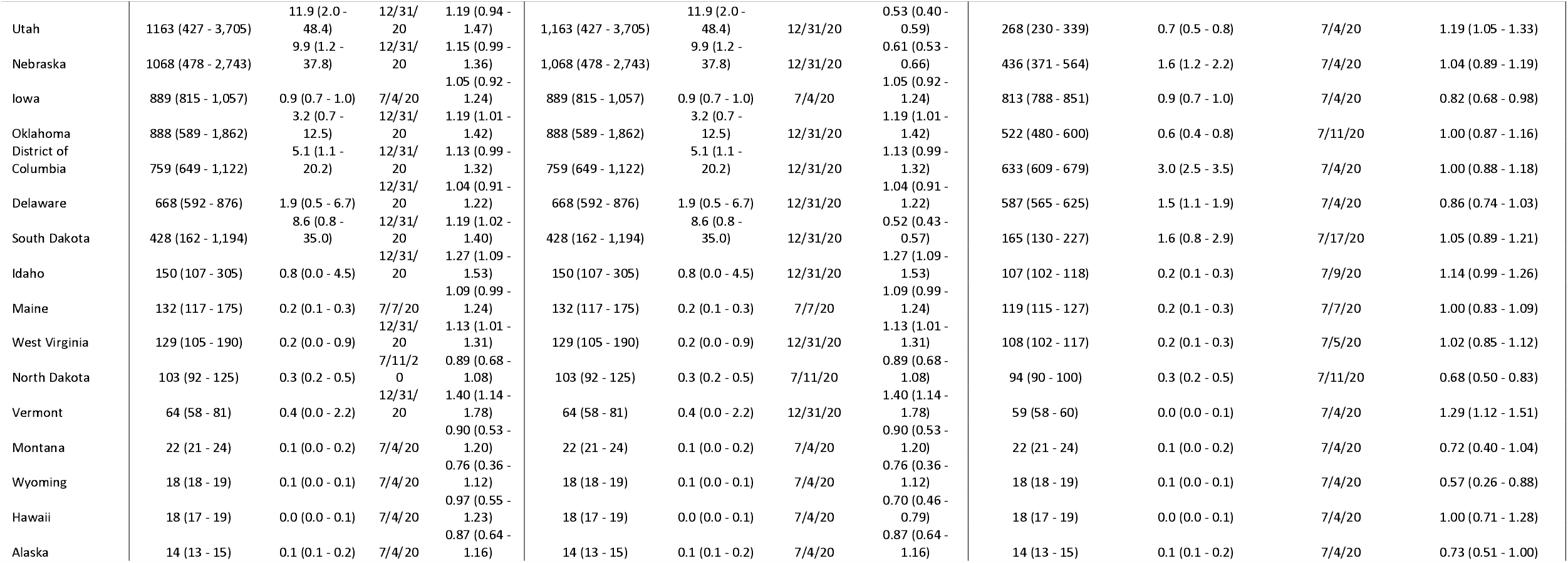
Cumulative deaths 04 July 2020 through 31 December 2020, maximum estimated daily deaths per million population, date of maximum daily deaths, and estimated R_effective_ on 31 December 2020 for three scenarios

When we model the future course of the epidemic assuming that states will move to once again shut down social interaction and economic activity when daily deaths reach a threshold of 8 deaths per million (the plausible “reference” scenario), the projected cumulative death toll across the US is forecast to be lower than under the “mandates easing” scenario, with 294,565 (233,885–398,397) deaths by 31 December 2020 (Fig. 2). Thus, across the 24 states that are projected to exceed 8 deaths per million under the “mandates easing” scenario by the end of 2020 (Table 1), the re-imposition of SDM could save 135,929 (49,669–278,666) lives. This scenario results in 30,336,701 (12,044,797–55,506,392) fewer estimated infections across the United States by the end of year (Extended Data Fig. 5) compared to the “mandates easing” scenario, with the highest rates of infections estimated to occur in New Jersey (24.9% (21.8–30.8%) infected), Massachusetts (21.2% (18.0–27.8%) infected), and Louisiana (19.4% (12.6–33.8%) infected) (Extended Data Fig. 6). As with the previous scenario, even with the re-imposition of SDM when daily deaths exceed 8 per million population, 47 states would reach an R_effective_ greater than one before the end of the year (Fig. 4, Table 1). Further results for hospital resource use needs are presented in Extended Data Figs 2,3 and forecast infections under this scenario are presented in Extended Data Figs 7,8.

The scenario where the population of each state was assumed to adopt and maintain the maximum observed level of mask use observed globally (see methods) – in addition to states re-imposing SDM if a threshold daily death rate of 8 deaths per million population was exceeded – resulted in the lowest projected cumulative death toll across US states, with a total of 191,771 (175,160– 223,377) deaths forecast to occur by 31 December 2020 (Fig. 2, Table 1). Under this scenario, at the time of the US national election on 3 November 2020, no states will have exceeded a daily death rate of 8 deaths per million (Fig. 3), although 38 states are still estimated to exceed an *R*_*effective*_ of one at some point between 4 July and 31 December 2020, and 33 states would have an *R*_*effective*_ greater than one on 31 December (Fig. 4). Through the end of the year, the daily death rate is forecast to exceed 8 deaths per million in just three states (California, Massachusetts, and Virginia) (Table 1) saving 102,795 (55,898–183,374) lives when compared to the plausible reference scenario and 238,723 (112,886– 426,205) lives when compared to the “mandates easing” scenario. Universal mask use combined with threshold-driven imposition of SDM results in 12,920,928 (7,136,980–22,826,322) fewer estimated infections across the United States by the end of year compared to the plausible reference scenario, and 43,257,629 (19,744,352–74,125,020) fewer estimated infections compared to the “mandates easing” scenario (Extended Data Fig. 5). The highest infection rates under the mask use scenario are estimated to occur in Massachusetts (21.0% (17.3–29.9%) infected), New Jersey (20.7% (19.3–22.5%) infected), and New York (17.8% (16.8–18.7%) infected) (Extended Data Fig. 6). Further results for hospital resource use needs are presented in Extended Data Figs 2,3 and forecast infections under this scenario are presented in Extended Data Figs 7,8.

## Discussion

We delimit three possible futures (continued removal of SDM, plausible reference, and universal mask-use scenarios), to help frame and inform a national discussion on what actions can be taken during the summer of 2020 and the profound public health, economic, and political influences these decisions will have for the rest of the year. Under all scenarios, the US is likely to face a continued public health challenge from the COVID-19 pandemic through December 2020 and beyond, with populous states in particular facing high levels of illness, deaths, and hospital demands from the disease. The implementation of SDMs as soon as individual states reach a threshold of 8 daily deaths per million can dramatically ameliorate the effects of the disease; achieving near universal mask use could delay or prevent this threshold from being reached in many states and has the potential to save the most lives while minimizing damage to the economy. National and state-level decision makers can use these forecasts of the potential health benefits of available NPI alongside considerations of economic and other social costs to make the most informed decisions on how to confront the COVID-19 pandemic at the local level. Our findings indicate that mask use, a relatively affordable and low-impact intervention, has the potential to serve as a priority life-saving strategy in all US locations.

New epidemics, resurgences, and second waves are not inevitable. Several countries have sustained reductions in COVID-19 cases over time^32^. Early indications that seasonality may play a role in transmission, with increased spread during colder winter months as is seen with other respiratory viruses^34–37^, highlight the importance of taking action both before and during the pneumonia season in the US. While it is yet unclear if COVID-19 seasonality will match that of pneumonia in general, the strong association observed so far should be heeded as a plausible warning of what is to come. Toward the end of 2020, masks could contain a second wave of resurgence while reducing the need for frequent and widespread imposition of SDMs. Such an approach has the potential to save lives while minimizing the economic and societal disruption associated with both restrictive SDMs and the pandemic itself. Although 95% mask use across the population may seem like a high threshold to achieve and maintain, this value represents a level that has been achieved elsewhere (see methods and Supplementary Information section 3.4). Where mask use has been widely adopted, in South Korea, Hong Kong, Japan, and Iceland, among others, transmission has declined and in some cases halted^32^. These examples serve as additional natural experiments^38^ of the likely impact of masks and support the findings from the universal mask use scenario. Long-term, the future of COVID-19 in the US will be determined by the evolution of herd immunity through progressive pandemic waves over seasons and/or through the deployment of an efficacious vaccine or therapeutic approaches.

Mask use has emerged as a contentious issue in the US. At the same time, although well below the rates seen in other countries, about 41% of US residents have reported that they “always” wear a mask^31^. The highest proportions of mask use were reported in the northeast of the country, where several states had estimated mask use greater than 60% on 26 June 2020^31^. The potential life-saving benefit of increasing mask use in the coming summer and fall cannot be overstated. Recent large-scale outdoor gatherings, such as the massive marches and protests against police brutality and racism that took place in June 2020 in the US, seem to have had a negligible effect on SARS-CoV-2 infection rates^39^ possibly due to high levels of mask use^40^. As Americans prepare to head to the polls in November, local policy makers should consider the health implications of long lines at polling places and the role of mask use (or alternatives such as mail-in voting) in mitigating disease spread. Several states have already postponed primary elections in an effort to avoid increased transmission. Mandatory mask laws have also been introduced in many states^38,41^, but compliance appears to be variable, indicating that mandates alone may be insufficient to substantially alter behavior. In certain locations, such as prisons, mask use alone may not be sufficient to prevent transmission, social distancing may not be feasible, and alternate solutions to protect these vulnerable populations may be needed^42^. Ultimately, US residents will need to choose between higher levels of mask use or risking the frequent redeployment of more stringent and economically damaging SDMs; or, in the absence of either measure, face a reality of a rising death toll^43^.

This work represents the outputs of a class of models that aim to abstract the disease transmission process in populations to a level that is tractable for understanding, and, in this case, that can be used for predictions. A clear consequence of any such exercise is that it will be limited by data (disease and relevant covariates), the model of understanding developed, and the length of time available to the model to learn/train the important dynamics. We have therefore tried to benchmark our model against alternative models of the COVID-19 pandemic and fully document our predictive performance with a range of measures^44^. In addition, we have provided the reader all the data and model code to enable full reproducibility and increased transparency and presented a range of likely futures in the form of a continued removal of mandates, plausible reference, and universal mask use scenario for decision makers to review. In addition, triangulation of other outputs of the SEIR model, such as the proportion of the population that are affected, are also provided and tested against independent data, in this case seroprevalence surveys (Extended Data Fig. 9). Finally, because uncertainty compounds with distance into the future predicted, the data, model, and its assumptions will be iteratively updated as the pandemic continues to unfold.

As we extend this work to investigate the impact of mask use and other NPI on the global pandemic, we are hopeful that masks will be sufficient in all states to avoid a COVID-19 resurgence in the US and avoid further economic damage. The US can reduce a potential second wave, if its residents decide to do so.

## Online content

Results for each state are accessible through a visualization tool at http://covid19.healthdata.org. The estimates presented in this tool will be iteratively updated as new data are incorporated and will ultimately supersede the results in this paper.

## Data Availability

All estimates can be further explored through our customized online data visualization tools (https://covid19.healthdata.org/united-states-of-america). The findings of this study are supported by data available in public online repositories, data publicly available upon request of the data provider, and data not publicly available owing to restrictions by the data provider. Non-publicly available data were used under license for the current study but may be available from the authors upon reasonable request and with permission of the data provider. Detailed tables and figures of data sources and availability can be found in SI Figures 1-4, and SI Tables 1-11. All maps presented in this study are generated by the authors using RStudio (R Version 3.6.3) and no permissions are required to publish them. Administrative boundaries were retrieved from the Database of Global Administrative Areas (GADM). Land cover was retrieved from the online Data Pool, courtesy of the NASA EOSDIS Land Processes Distributed Active Archive Center (LP DAAC), USGS/Earth Resources Observation and Science (EROS) Center, Sioux Falls, South Dakota. Populations were retrieved from WorldPop (https://www.worldpop.org).

## Methods

Our analysis strategy supports two main and interconnected objectives: (1) generate predictions of COVID-19 deaths, infections, and hospital resource needs for all US states; and (2) explore alternative scenarios on the basis of changes in state-imposed social distancing mandates or population levels of mask use. The modeling approach to achieve this is summarized in Supplementary Information section 2 and can be divided into four stages: (1) identification and processing of COVID-19 data, (2) exploration and selection of key drivers or covariates, (3) modelling deaths and cases across three scenarios of SDM in US states using an SEIR framework, and (4) modeling heath service utilization as a function of forecast infections and deaths within those scenarios. This study complies with the Guidelines for Accurate and Transparent Health Estimates Reporting (GATHER) statement (Supplementary Information).

### Data identification and processing

IHME forecasts include data from local and national governments, hospital networks and associations, the World Health Organization, third-party aggregators, and a range of other sources. Data sources and corrections are described in detail in the Supplementary Information. Briefly, daily confirmed case and death numbers due to COVID-19 are collated from the Johns Hopkins University (JHU) data repository; we supplement and correct this dataset as needed to improve the accuracy of our projections and adjust for reporting-day biases (see Supplementary Information Table 4). Testing data are obtained from the *Our World in Data* COVID tracking project and supplemented with data from additional government websites (Supplementary Information Table 8). Social distancing data are obtained from a number of different official and open sources, which vary by state (Supplementary Information Table 7). Mobility data are obtained from Facebook Data for Good, Google, SafeGraph, and Descartes Labs (Supplementary Information section 3.2). Mask use data are obtained from the Facebook Global Symptom Survey (in collaboration with the University of Maryland Social Data Science Center) and PREMISE (Supplementary Information section 3.4). Specific sources for data on licensed bed and ICU capacity and average annual utilization in the United States are detailed in the Supplementary Information section 2.

Before modeling, observed cumulative deaths are smoothed using a spline-based smoothing algorithm with randomly placed knots. Uncertainty is derived from bootstrapping and resampling of the observed deaths. The time series of case data is used as a leading indicator of death based on an infection fatality ratio (IFR) and a lag from infection to death. These smoothed estimates of observed deaths by location are then used to create estimated infections based on an age-distribution of infections and on age-specific IFRs. The age-specific infections were collapsed into total infections by day and state and used as data inputs in the SEIR model. Detailed descriptions of data smoothing and transformation steps are provided in the Supplementary Information.

### Covariate selection

Covariates for the compartmental transmission SEIR model are predictors of the *β* parameter in the model that affects the transition from Susceptible to Exposed state. Covariates were evaluated on the basis of biologic plausibility and on the impact on the results of the SEIR model. Given limited empirical evidence of population-level predictors of SARS-CoV-2 transmission, biologically plausible predictors of pneumonia such as population density (percentage of the population living in areas with more than 1000 individuals per square kilometer), tobacco smoking prevalence, population-weighted elevation, lower respiratory infection mortality rate, and particulate matter air pollution were considered. These covariates are representative at a population level and are time-invariant. Spatially resolved estimates for these covariates are derived from the Global Burden of Disease Study 2019^45^. Time-varying covariates include pneumonia excess mortality seasonality, diagnostic tests per capita, population-level mobility, and personal mask use. These are described in the following sections.

### Pneumonia seasonality

We used weekly pneumonia mortality data from the National Center for Health Statistics Mortality Surveillance System^46^ from 2013 to 2019 by US state. Pneumonia deaths included all deaths classified by the full range of ICD codes in J12–J18.9. We pooled data over available years for each state and found the weekly deviation from the annual, state-specific mean mortality due to pneumonia. We then fit a seasonal pattern using a Bayesian meta-regression model with a flexible spline and assumed annual periodicity (Supplementary Information section 3.5). For locations outside the United States, we used vital registration data where available. Locations without vital registration data had weekly pneumonia seasonality predicted based on latitude from a model pooling all available data (Supplementary Information section 3.5).

### Testing *per capita*

We considered diagnostic testing for active SARS-CoV-2 infections as a predictor of the ability for a state to identify and isolate active infections. We assumed that higher rates of testing are negatively associated with SARS-CoV-2 transmission. Our primary sources for US testing data were compiled by the COVID Tracking Project (Supplementary Information section 3.3 and SI Table 8). Unless testing data existed before the first confirmed case in a state, we assumed that testing is non-zero after the date of the first confirmed case. Before producing predictions of testing per capita, we smoothed the input data by using the same smoothing algorithm used for smoothing daily death data prior to modeling (previously described). Testing per capita projections for unobserved future days were based on linearly extrapolating the mean day-over-day difference in daily tests per capita for each location. We put an upper limit on diagnostic tests per capita of 500 per 100,000 based on the highest observed rates in June 2020.

### Social distancing mandates

Social distancing mandates (SDMs) were not used as direct covariates in the transmission model. Rather, SDMs were used to predict population mobility (see below) which is subsequently used as a covariate in the transmission model. We collected the dates of state-issued mandates enforcing social distancing as well as the planned or actual removal of these mandates. The measures that we included in our model were 1) severe travel restrictions, 2) closing of public educational facilities, 3) closure of non-essential businesses, 4) stay at home orders, 5) restrictions on gathering size. Generally, these came from state government official orders or press releases.

To determine the expected change in mobility due to social distancing mandates, we used a Bayesian, hierarchical meta-regression model with random effects by location on the composite mobility indicator to estimate the effects of social distancing policies on changes in mobility (Supplementary Information section 3.1).

### Mobility

We used four data sources on human mobility to construct a composite mobility indicator. Those sources were Facebook, Google, SafeGraph, and Descartes Labs (Supplementary Information section 3.2). Each source has a slightly different way of capturing mobility, so before constructing a composite mobility indicator, we standardized these different data sources (Supplementary Information section 3.2). Briefly, this first involved determining the change in a baseline level of mobility for each location by data source. Then, we determined a location-specific median ratio of change in mobility for each pairwise comparison of mobility sources, using Google as a reference and adjusting the other sources by that ratio. The time series for mobility was estimated using a Gaussian process regression model using the standardized data sources to get a composite indicator for change in mobility for each location-day.

We calculated the residuals between our predicted composite mobility time series and input composite time series, and then applied a first-order random walk to the residuals. The random walk was used to predict residuals from 01 January 2020 to 01 January 2021, which were then added to the mobility predictions to produce a final time series with uncertainty: “past” changes in mobility from 01 January 2020 to 27 June 2020, and projected mobility from 27 June 2020 to 01 January 2021.

### Masks

We performed a meta-analysis of 40 peer-reviewed scientific studies in an assessment of mask effectiveness for preventing respiratory viral infections (Supplementary Information section 3.4). The studies were extracted from a preprint publication^28^. In addition, we considered all articles from a second meta-analysis^30^ and one supplemental publication^47^. These studies included both persons working in health care and the general population – especially family members of those with known infections. The studies indicate overall reductions in infections due to masks preventing exhalation of respiratory droplets containing viruses, as well as some prevention of inhalation by those uninfected. The resulting meta-regression calculated log-transformed relative risks and corresponding log-transformed standard errors based on raw counts and used a continuity correction for studies with zero counts in the raw data (0.001). Whereas the other meta-analyses reported one outcome per study, we extracted all relevant outcomes per study. Additionally, we included additional specifications and characteristics to account for differences in characteristics of individual studies and to identify important factors impacting mask effectiveness. These include the type of population using masks (general population versus health care population), country of study (Asian countries versus non-Asian countries), type of mask (paper, cloth, or non-descript masks versus medical masks and N95 masks), type of control group (no use versus infrequent use), type of disease (SARS-CoV 1 or 2 versus H1N1, influenza, or other respiratory pathogens), and type of diagnosis (clinical versus laboratory).

We used MR-BRT – a meta-regression tool developed at the Institute for Health Metrics and Evaluation (meta-regression, Bayesian, regularized, trimmed) (Supplementary Information section 2.5) – to perform a meta-analysis that considered the various characteristics of each study. We accounted for between-study heterogeneity and quantified remaining between-study heterogeneity into the width of the uncertainty interval. We also performed various sensitivity analyses to verify the robustness of the modeled estimates and found that the estimate of the effectiveness of mask use did not change significantly when we explored four alternative analyses, including changing the continuity correction assumption, using odds ratio versus relative risk from published studies, using a fixed effects versus a mixed effects model, and including studies without covariate information.

We estimated the proportion of people who self-reported always wearing a face mask when outside in public for both US and global locations using data from PREMISE (US) and Facebook (non-US). We again used the same smoothing model as for COVID-19 deaths and testing per capita to produce estimates of observed mask use. This smoothing process averaged each data point with its neighbors. Tails are an average of the change in mask use over the three following days (left tail) and three preceding days (right tail). The level of mask use starting on 26 June 2020 (or the last day of processed and analyzed data) is assumed to be flat. Among states without state-specific data, a regional average was used.

### Deterministic modeling framework

Model specification is provided in detail in the Supplementary Information and summarized in a schematic (SI Fig. 1). In order to fit and predict disease transmission dynamics, we include a susceptible-exposed-infected-recovered (SEIR) component in our multi-stage model. In particular, each location’s population is tracked through the following system of differential equations:

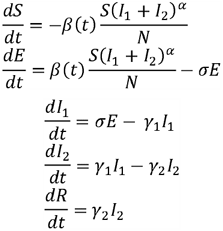

where *α* represents a mixing coefficient to account for imperfect mixing within each location, *σ* is the rate at which infected individuals become infectious, *γ*_1_ is the rate at which infectious people transition out of the pre-symptomatic phase, and *γ*_2_ is the rate at which individuals recover. This model does not distinguish between symptomatic and asymptomatic infections but has two infectious compartments (*I*_l_ and *I*_2_) to allow for interventions that would avoid focus on those who could not be symptomatic; *I*_l_ is thus the pre-symptomatic compartment.

Using the next-generation matrix approach, we can directly calculate both the basic reproductive number under control (*R*_c_(t)) and the effective reproductive number (*R*_*effective*_ (t)) as (see Supplementary Information section 5.1 for derivation):

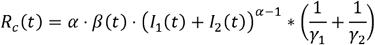

and

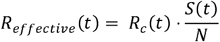

By allowing *β* (*t*) to vary in time, our model is able to account for increases in transmission intensity as human behavior shifts over time (e.g., changes in mobility, adding or removing SDM, changes in population mask use). Briefly, we combine data on cases (correcting for trends in testing), hospitalizations, and deaths into a distribution of trends in daily deaths.

To fit this model, we resample 1000 draws of daily deaths from this distribution for each state (see Supplementary Information section 5). Using an estimated IFR by age (Supplementary Information section 4.2) and the distribution of time from infection to death (Supplementary Information section 4.3), we then use the daily deaths to generate 1000 distributions of estimated infections by day from 10 January to 04 July 2020. We then fit the rates at which infectious individuals may come into contact and infect susceptible individuals (denoted as *β* (*t*)) as a function of a number of predictors that affect transmission. Our modeling approach acts across the overall population (i.e., no assumed age structure for transmission dynamics), and each location is modeled independently of the others (i.e., we do not account for potential movement between locations).

We detail the SEIR fitting algorithm in the Supplementary Information section 5.1, but in brief, by draw we first fit a smooth curve to our estimates of daily new infections. Then, sampling *γ*_2_, *σ*, and *α* from defined ranges from literature (see SI) and using 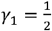 we then sequentially fit the *E,I*_1_,*I*_2_, and *R* components in the past. We then algebraically solve the above system of differential equations for *β* (*t*).

The next stage of our model fits relationships between past changes in *β* (*t*) and covariates described above: mobility, testing, masks, pneumonia seasonality, others. As detailed in Supplementary Information section 3, the time-varying covariates are forecast from 01 July to 31 December 2020. The fitted regression is then used to estimate future transmission intensity *β*_*pred*_ The final future transmission intensity is then an adjusted version of *β*_*pred*_ based on the average fit over the recent past (where the window of averaging varies by draw from 2 to 4 weeks; see Supplementary Information section 5 for more details).

Finally, we use the future estimated transmission intensity to predict future transmission (using, for each draw, the same parameter values for all other SEIR parameters). In a reversal of the translation of deaths into infections, we then use the estimated daily new infections to calculate estimated daily deaths (again using the location-specific IFR). We also use the estimated trajectories of each SEIR compartment to calculate *R*_c_ and *R*_*effective*_.

A final step to take predicted infections and deaths and a hospital use microsimulation to estimate hospital resource need for each US state is described in greater detail in the Supplementary Information section 7 and the results presented online (https://covid19.healthdata.org/united-states-of-america).

### Forecasts/scenarios

Policy responses to COVID-19 can be supported by the evaluation of impacts of various scenarios of those options, against a background of business as usual assumption, to explore fully the potential impact of policy levers available.

We estimate the trajectory of the epidemic by state under a “mandates easing” scenario that models what would happen in each state if the current pattern of easing social distancing mandates continues and new mandates are not imposed. This should be thought of as a worst-case scenario, where regardless of how high the daily death rate gets, SDM will not be re-introduced and behavior (including population mobility and mask use) will not vary before 31 December 2020. In locations where the number of cases is rising, this leads to very high predictions by the end of the year.

As a more plausible scenario, we use the observed experience from the first phase of the pandemic to predict the likely response of state and local governments during the second phase. This plausible reference scenario assumes that in each location the trend of easing SDM will continue at its current trajectory until the daily death rate reaches a threshold of 8 deaths per million. If the daily death rate in a location exceeds that threshold, we assume that SDM will be reintroduced for a six-week period. The choice of threshold (of a rate of daily deaths of 8 per million) represents the 90^th^ percentile of the distribution of daily death rate at which US states implemented their mandates during the first months of the COVID-19 pandemic. We selected the 90^th^ percentile rather than the 50^th^ percentile to capture an anticipated increased reluctance from governments to re-impose mandates because of the economic effects of the first set of mandates. In locations that do not exceed the threshold of a daily death rate of 8 per million, the projection is based on the covariates in model and the forecasts for these to 31 December 2020. In locations were the daily death rate exceeded 8 per million at the time of our final model run for this manuscript (04 July 2020), we are assuming that mandates will be introduced within seven days.

The scenario of universal mask wearing models what would happen if 95% of the population in each state always wore a mask when they were in public. This value was chosen to represent the highest observed rate of mask use in the world so far during the COVID-19 pandemic (see Supplementary Information section 3.4). In this scenario, we also assume that if the daily death rate in a state exceeds 8 deaths per million, SDMs will be reintroduced for a six-week period.

### Model validation

Model performance was tested against reported deaths from 01 February to 30 June 2020^24^. Out-of-sample predictive validity was assessed periodically for all model versions against subsequently observed trends in COVID-19 weekly and cumulative mortality. The IHME hybrid SEIR model described here was found to have a median absolute percent error of 9.9% at four weeks after the last available input data^25^. This work provides a comprehensive and reproducible platform for testing model performance for the model presented here and all other models that have published and archived similar predictions.

The increasing number of population-based serology surveys conducted also provide a unique opportunity to cross-validate our prior predictions with modeled epidemiological outcomes. In Extended Data Fig. 9 we compare these serology surveys (such as the Spanish ENE-COVID study^48^) to our estimated population seropositivity time-indexed to the date that the survey was conducted. In general, across the varied locations that have been reported globally, we note a high degree of agreement between the estimated and surveyed seropositivity. As more serology studies are conducted and published, especially in the US, this will allow an ongoing and iterative assessment of model validity.

## Limitations

Epidemics progress based on complex non-linear and dynamic biological and social processes that are difficult to observe directly and at scale. Mechanistic models of epidemics, formulated either as ordinary differential equations or as individual-based simulation models, are a useful tool for conceptualizing, analyzing, or forecasting the time course of epidemics. In the COVID-19 epidemic, effective policies and the responses to those policies have changed the conditions supporting transmission from one week to the next, with the effects of policies realized typically after a variable time lag. Each model approximates an epidemic, and whether used to understand, forecast, or advise, there are limitations on the quality and availability of the data used to inform it and the simplifications chosen in model specification. It is unreasonable to expect any model to do everything well, so each model makes compromises to serve a purpose, while maintaining computational tractability.

One of the largest determinants of the quality of a model is the corresponding quality of the input data. Our model is anchored to daily COVID-19-related deaths, as opposed to daily COVID-19 case counts, due to the assumption that death counts are a less biased estimate of true COVID-19-related deaths than COVID-19 case counts are of the true number of SARS-CoV-2 infections. Numerous biases such as treatment-seeking behavior, testing protocols (such as only testing those who have traveled abroad), and differential access to care greatly influence the utility of case count data. Moreover, there is growing evidence that inapparent and asymptomatic individuals are infectious as well as individuals who eventually become symptomatic being infectious before the onset of any symptoms. As such, our primary input data for our model are counts of deaths; death data can likewise be fallible, however, and where available, we combine death data, case data, and hospitalization data together to estimate COVID-19 deaths.

Beyond the basic input data, there are a large number of other data sources with their own potential biases that are incorporated into our model. Testing, mobility, and mask use are all imperfectly measured and may or may not be representative of the practices of those that are susceptible and/or infectious. Moreover, any forecast of the patterns of these covariates is associated with a large number of assumptions (detailed in the corresponding sections of the Supplementary Information), and as such, care must be taken in the interpretation of estimates farther into the future, as the uncertainty associated with the numerous sub-models that go into these estimates increases in time.

For practical purposes, our transmission model has made a large number of simplifying assumptions. Key among these is the exclusion of movement between locations (e.g., importation) and the absence of age structure and mixing within location (e.g., we assume a well-mixed population). It is clear that there are large, super-spreader-like events that have occurred throughout the COVID-19 pandemic, and our current model is unable to fully capture these dynamics within our predictions. Another important assumption to note is that of the relationship between pneumonia seasonality and SARS-CoV-2 seasonality. To date, across both the northern and southern hemisphere, there is a strong association between COVID-19 cases and deaths and general seasonal patterns of pneumonia deaths (SI Section 3.5). Our predictions through the end of 2020 are immensely influenced by the assumption that this relationship will maintain through the year and that SARS-CoV-2 seasonality will be well approximated by pneumonia seasonality. While we assess this assumption to the extent possible (see Supplementary Information), we have not yet experienced a full year of SARS-CoV-2 transmission, and as such cannot yet know if this assumption is valid.

Finally, the model presented herein is not the first model our team has developed to predict current and future transmission of SARS-CoV-2. As the outbreak has progressed, we have attempted to adapt our modeling framework to both the changing epidemiological landscape as well as the increase in data that could be useful to inform a model^49^. Changes in the dynamics of the outbreak overwhelmed both the initial purpose and some key assumptions of our first model, requiring evolution in our approach. While the current SEIR formulation is a more flexible framework (and thus less likely to need to be wholly reconfigured as the outbreak progresses further), we fully expect the need to adapt our model to accommodate future shifts in patterns of SARS-CoV-2 transmission. Incorporating movement within and without locations is one example, but resolving our model at finer spatial scales as well as accounting for differential exposure and treatment rates across sexes and races are other dimensions of transmission modelling we currently do not account for but expect will be necessary additions in the coming months. As we have done before, we will continually adapt, update, and improve our model based on need and predictive validity.

## Code availability statement

Our study follows the Guidelines for Accurate and Transparent Health Estimate Reporting (GATHER; Supplementary Information). All code used for these analyses is publicly available online (http://github.com/ihmeuw/).

## Acknowledgments

We thank the various Departments of Health and frontline health professionals who are not only responding to this epidemic daily, but also provide the necessary data to inform this work – IHME wishes to warmly acknowledge the support of these and others (http://www.healthdata.org/covid/acknowledgements) who have made our COVID-19 estimation efforts possible. This work was supported by the Bill & Melinda Gates Foundation, as well as funding from the state of Washington and the National Science Foundation (2031096). We also extend a note of particular thanks to John Stanton and Julie Nordstrom for their generous support.

## Additional information

Supplementary Information is available for this paper: Supplementary Text on data and methods, Supplementary Model descriptions, Supplementary References, Supplementary Figures 1-4, and Supplementary Tables 1-11.

## Extended Data Figure Legends

**EDF 1.**
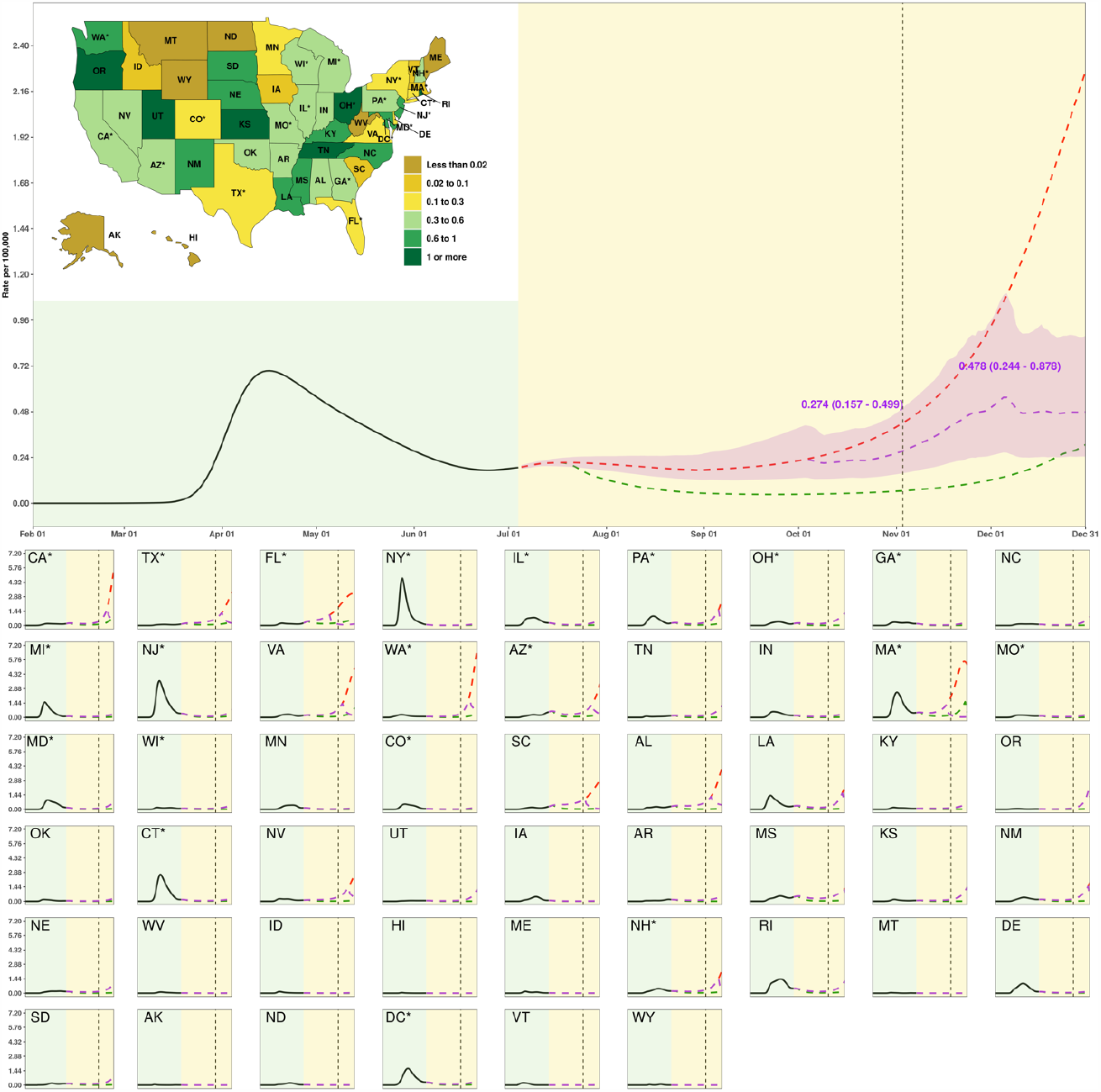
Estimated daily COVID-19 death rate (per 100,000 population) by state for three scenarios. The inset map displays the estimated peak in daily deaths from COVID-19 death per 100,000 population by state between 04 July and 31 December. The light yellow background separates the observed and predicted part of the time series, before and after 04 July. The dashed vertical line identifies 03 November 2020. The red line is the “mandates easing” scenario, the purple line the “plausible reference” scenario, and the green line the “universal mask” scenario. Numbers are the means and uncertainty interval (UI) for the plausible reference scenario on dates highlighted. State panels are ordered by decreasing population size. Two-letter state abbreviations are provided in panels and the inset map. An asterisk next to state abbreviation indicates a state with one or more urban agglomerations exceeding two million persons. State panels are scaled to accommodate the state with the highest value (WA here), ranging from zero to 7.2. This map was generated with RStudio (R Version 3.6.3).

**EDF 2.**
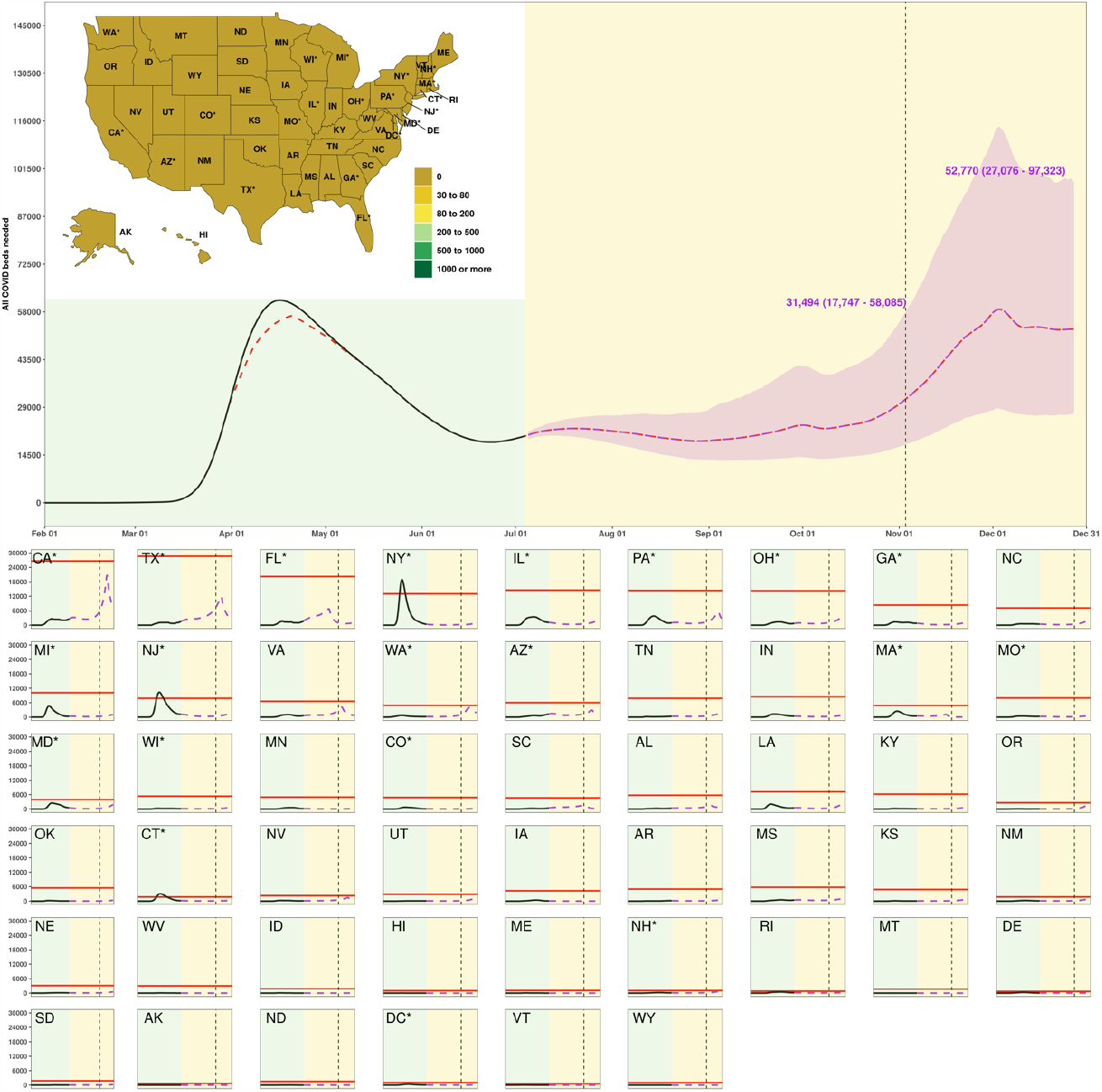
Estimated total hospital beds needed for COVID-19 patients by state from 01 February to 31 December 2020 for three scenarios. The inset map displays the estimated peak number of all COVID-19 beds above capacity by state between 04 July and 31 December. The light yellow background separates the observed and predicted part of the time series, before and after 04 July. The dashed vertical line identifies 03 November 2020. The purple line shows the time trend in estimated total hospital beds needed for COVID-19 patients under the “plausible reference” scenario; the horizontal red line identifies estimated total COVID-19 bed capacity for each state. Numbers are the mean and uncertainty interval (UI) for the plausible reference scenario on dates highlighted. State panels are ordered by decreasing population size. Two-letter state abbreviations are provided in panels and the inset map. An asterisk next to state abbreviation indicates a state with one or more urban agglomerations exceeding two million persons. State panels are scaled to accommodate the state with the most available all COVID beds (TX here), ranging from zero to 30,000. This map was generated with RStudio (R Version 3.6.3).

**EDF 3.**
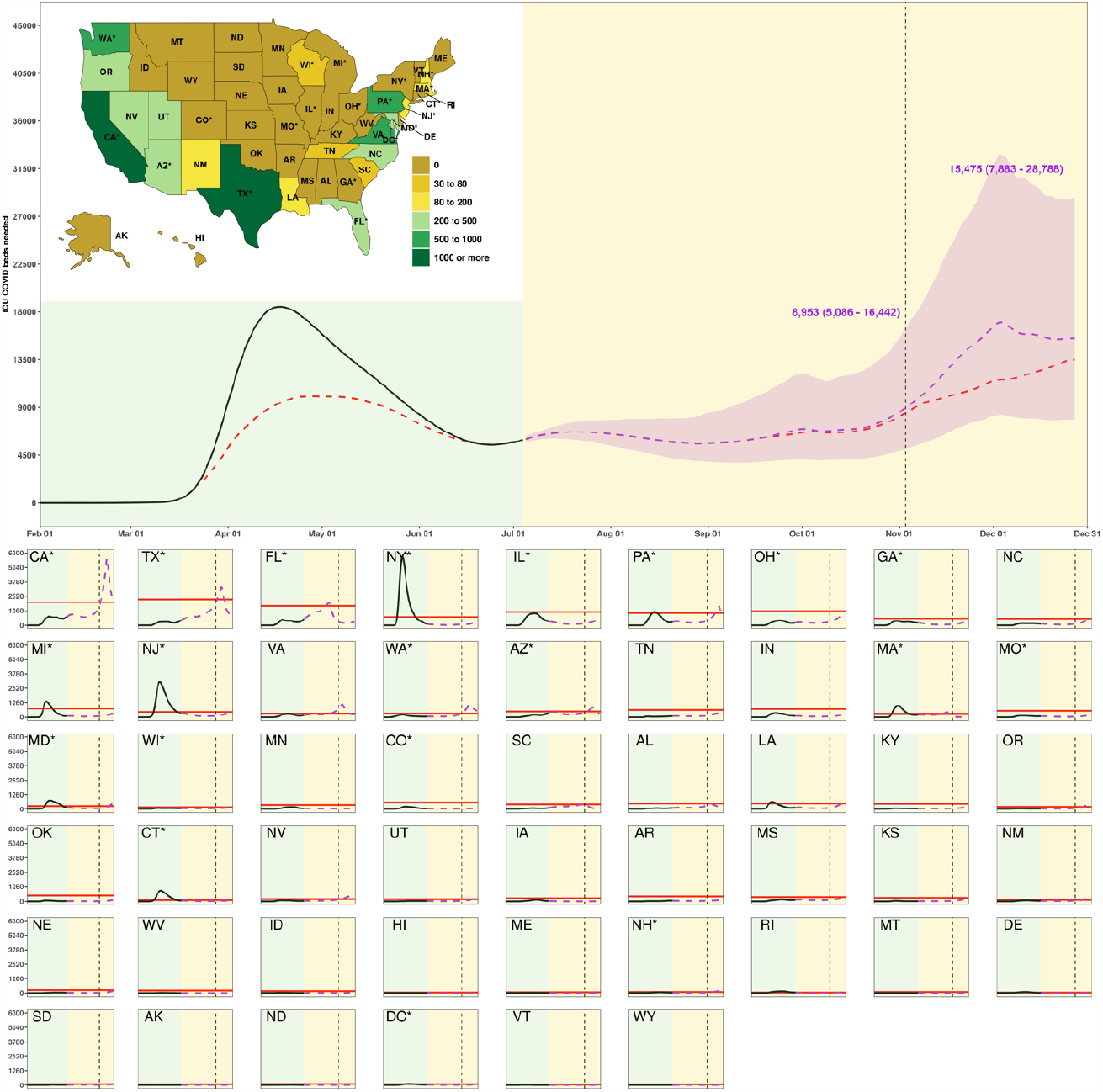
Estimated total ICU beds needed for COVID-19 patients by state from 01 February to 31 December 2020 for three scenarios. The inset map displays the estimated peak number of all ICU COVID-19 beds above capacity by state between 04 July and 31 December. The light yellow background separates the observed and predicted part of the time series, before and after 04 July. The dashed vertical line identifies 03 November 2020. The purple line shows the time trend in estimated total ICU beds needed for COVID-19 patients under the “plausible reference” scenario; the horizontal red line identifies estimated COVID-19 ICU bed capacity for each state. Numbers are the mean and uncertainty interval (UI) for the plausible reference scenario on dates highlighted. State panels are ordered by decreasing population size. Two-letter state abbreviations are provided in panels and the inset map. An asterisk next to state abbreviation indicates a state with one or more urban agglomerations exceeding two million persons. State panels are scaled to accommodate the state with the most ICU COVID beds needed (NY here), ranging from zero to 6,300. This map was generated with RStudio (R Version 3.6.3).

**EDF 4.**
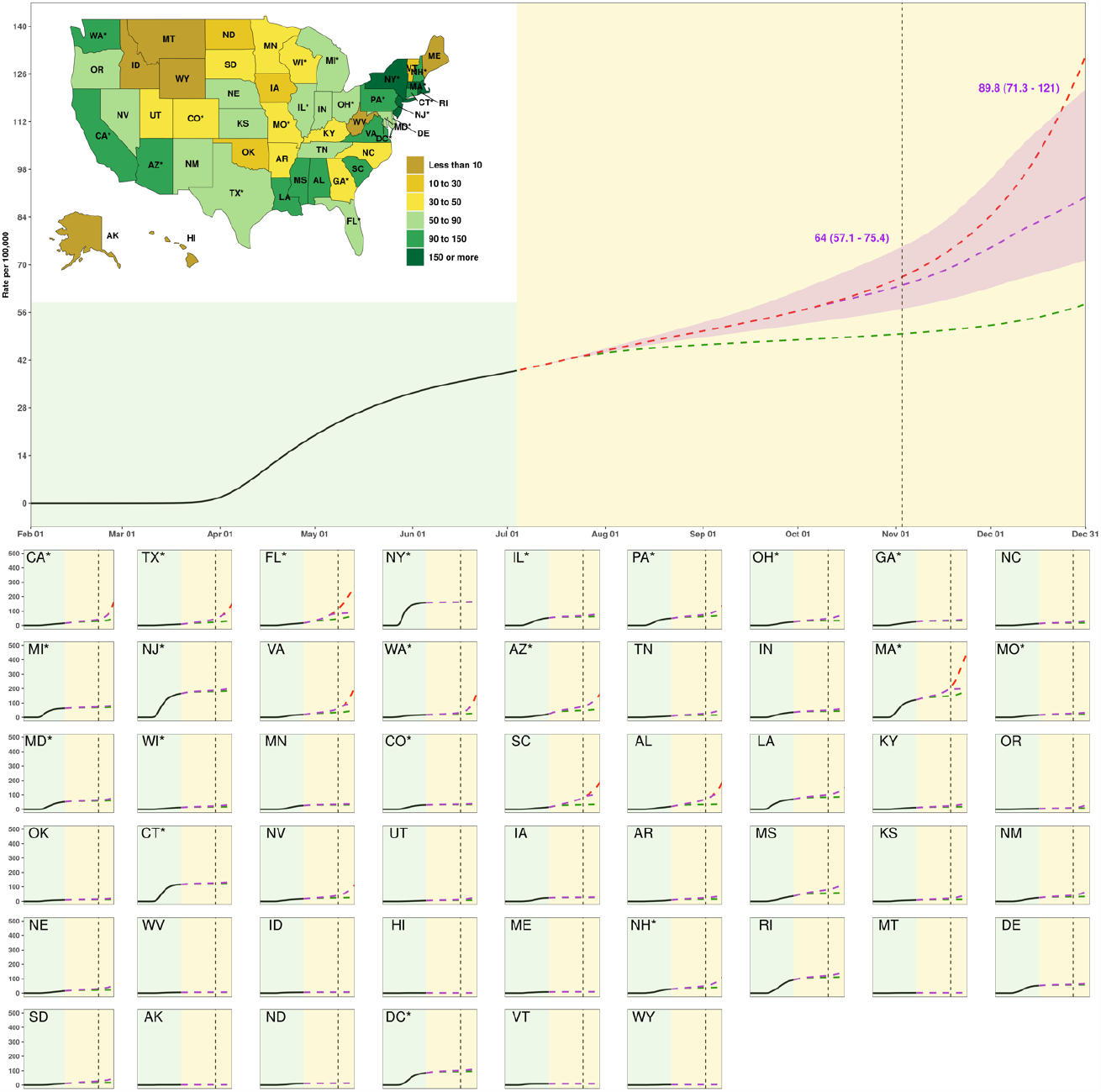
Estimated cumulative deaths from COVID-19 per 100,000 population from 01 February to 31 December 2020 by state for three scenarios. The inset map displays the cumulative deaths under the “plausible reference” scenario on 31 December 2020. The light yellow background separates the observed and predicted part of the time series, before and after 04 July. The dashed vertical line identifies 03 November 2020. The red line represents the estimated time trend for deaths in the “mandates easing” scenario, the purple line the “plausible reference” scenario, and the green line the “universal mask” scenario. Numbers are the mean and uncertainty interval (UI) for the plausible reference scenario on dates highlighted. State panels are ordered by decreasing population size. Two-letter state abbreviations are provided in panels and the inset map. An asterisk next to state abbreviation indicates a state with one or more urban agglomerations exceeding two million persons. State panels are scaled to accommodate the state with the highest value (MA here), ranging from zero to 500 deaths per 100,000. This map was generated with RStudio (R Version 3.6.3).

**EDF 5.**
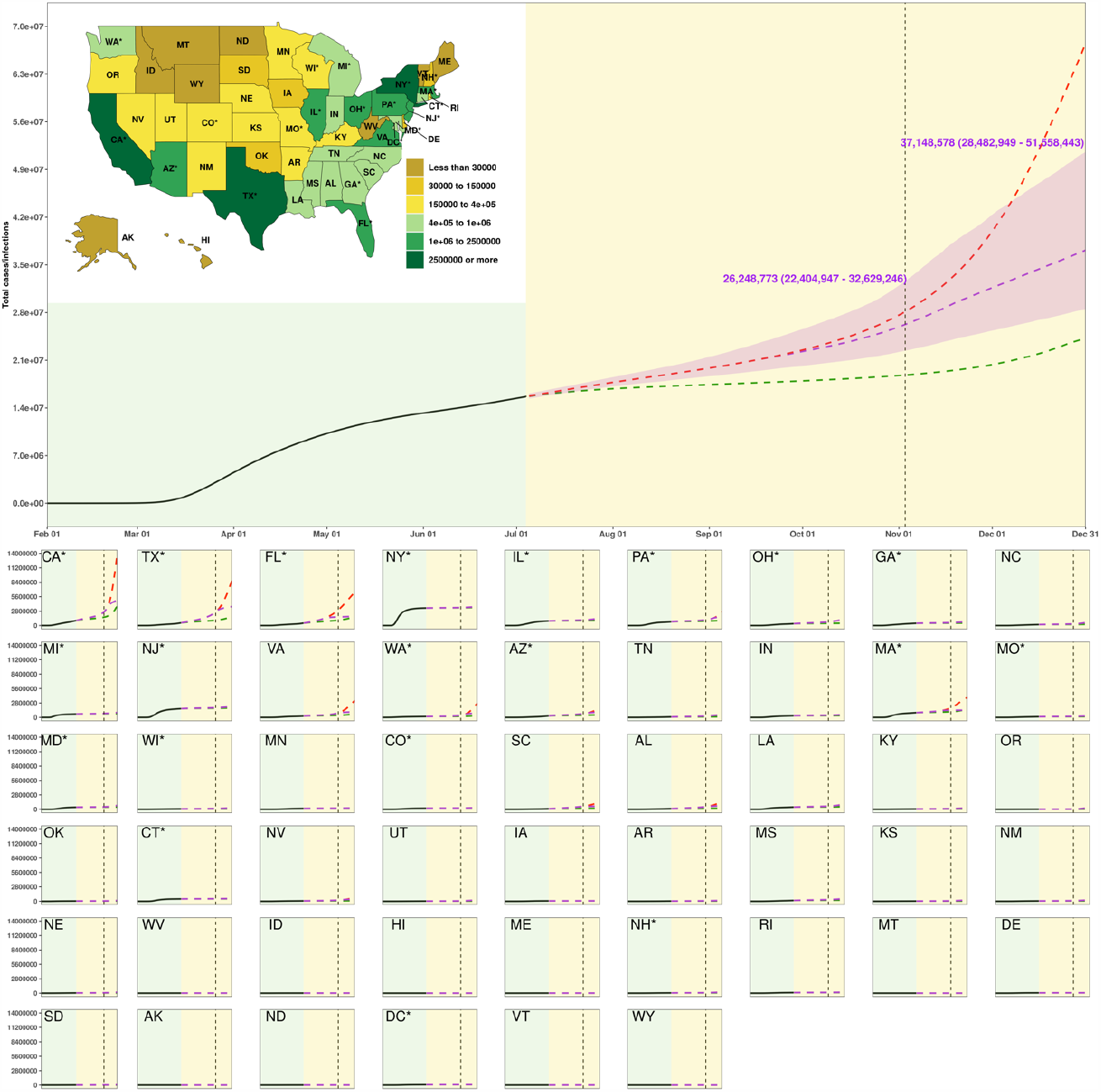
Estimated cumulative infections from SARS-CoV-2 from 01 February to 31 December 2020 by state for three scenarios. The inset map displays the cumulative infections under the “plausible reference” scenario on 31 December 2020. The light yellow background separates the observed and predicted part of the time series, before and after 04 July. The dashed vertical line identifies 03 November 2020. The red line represents the estimated time trend for infections in the “mandates easing” scenario, the purple line the “plausible reference” scenario, and the green line the “universal mask” scenario. Numbers are the mean and uncertainty interval (UI) for the plausible reference scenario on dates highlighted. State panels are ordered by decreasing population size. Two-letter state abbreviations are provided in panels and the inset map. An asterisk next to state abbreviation indicates a state with one or more urban agglomerations exceeding two million persons. State panels are scaled to accommodate the state with the highest value (CA here), ranging from zero to 14,000,000. This map was generated with RStudio (R Version 3.6.3).

**EDF 6.**
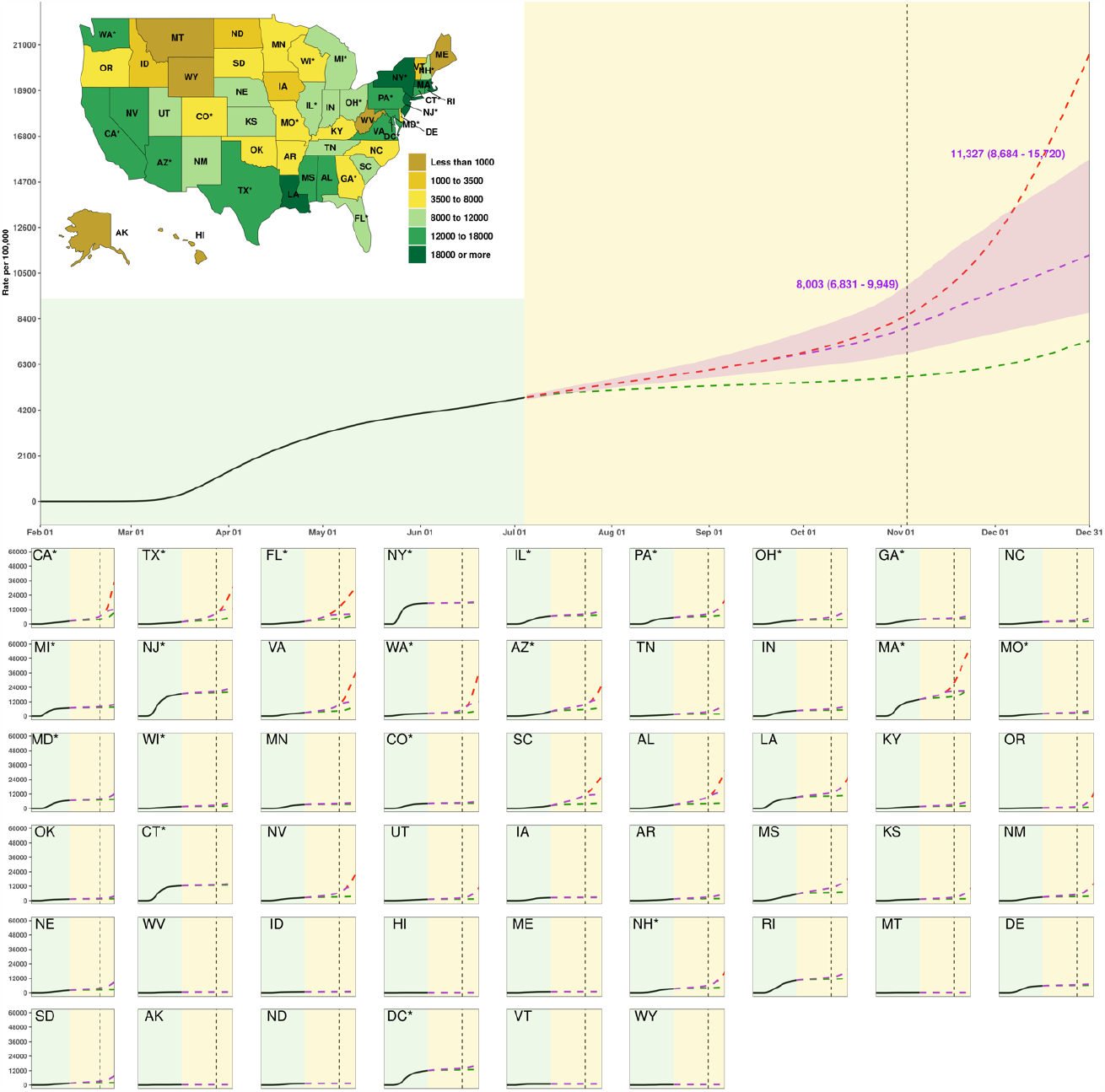
Estimated cumulative SARS-CoV-2 infection rate (per 100,000 population) by state for three scenarios. The inset map displays the estimated peak in cumulative infections from COVID-19 per 100,000 population by state between 04 July and 31 December 31. The light yellow background separates the observed and predicted part of the time series, before and after 04 July. The dashed vertical line identifies 03 November 2020. The red line is the “mandates easing” scenario, the purple line the “plausible reference” scenario, and green line the “universal mask” scenario. Numbers are the means and uncertainty interval (UI) for the plausible reference scenario on dates highlighted. State panels are ordered by decreasing population size. Two-letter state abbreviations are provided in panels and the inset map. An asterisk next to state abbreviation indicates a state with one or more urban agglomerations exceeding two million persons. State panels are scaled to accommodate the state with the highest value (MA here), ranging from zero to 60,000. This map was generated with RStudio (R Version 3.6.3).

**EDF 7.**
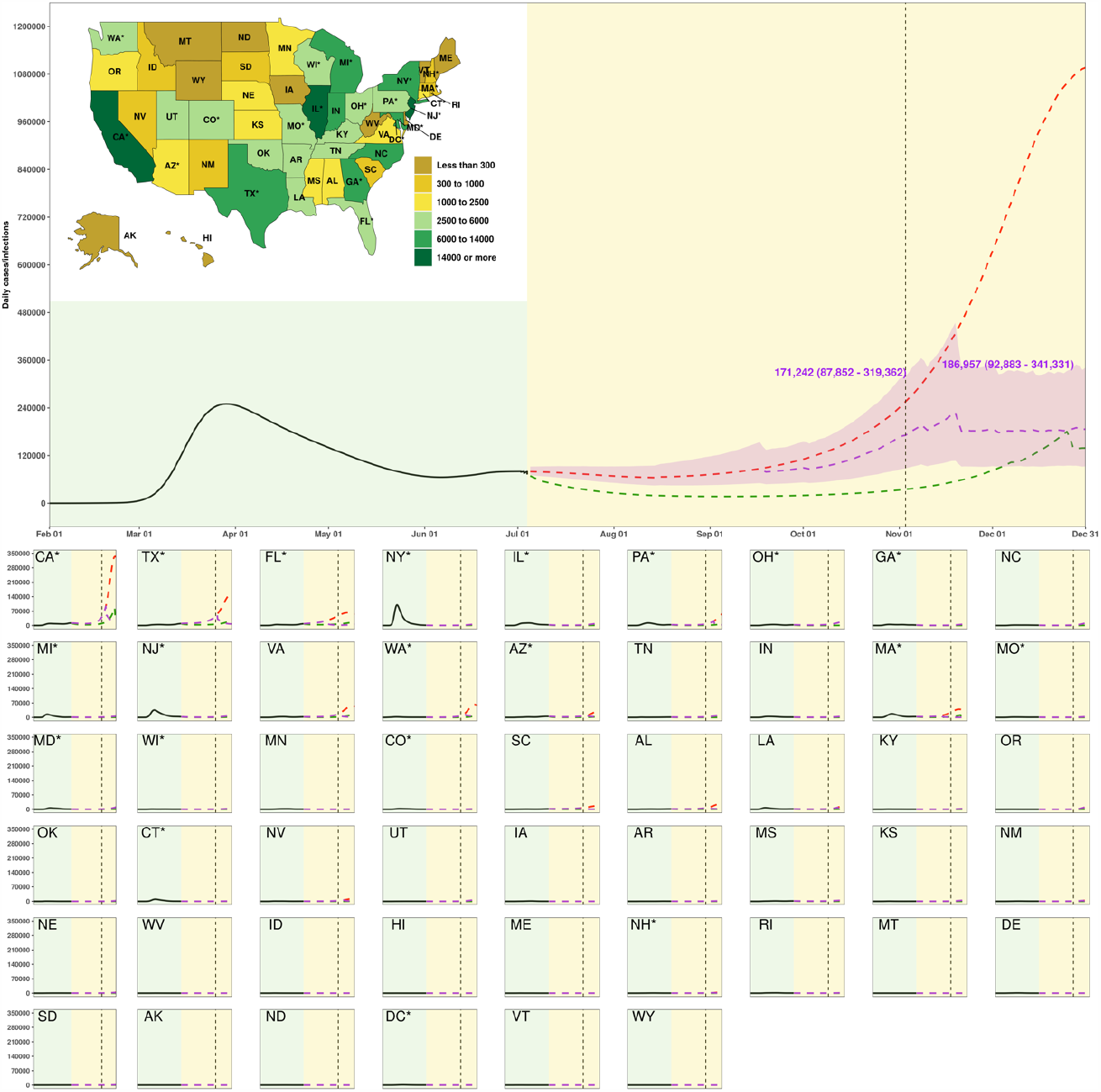
Estimated daily infections from SARS-CoV-2 from 01 February to 31 December 2020 by state for three scenarios. The inset map displays the daily infections under the “plausible reference” scenario on 31 December 2020. The light yellow background separates the observed and predicted part of the time series, before and after 04 July. The dashed vertical line identifies 03 November 2020. The red line represents the estimated time trend for daily infections in the “mandates easing” scenario, the purple line the “plausible reference” scenario, and the green line the “universal mask” scenario. Numbers are the mean and uncertainty interval (UI) for the plausible reference scenario on dates highlighted. State panels are ordered by decreasing population size. Two-letter state abbreviations are provided in panels and the inset map. An asterisk next to state abbreviation indicates a state with one or more urban agglomerations exceeding two million persons. State panels are scaled to accommodate the state with the highest value (CA here), ranging from zero to 350,000. This map was generated with RStudio (R Version 3.6.3).

**EDF 8.**
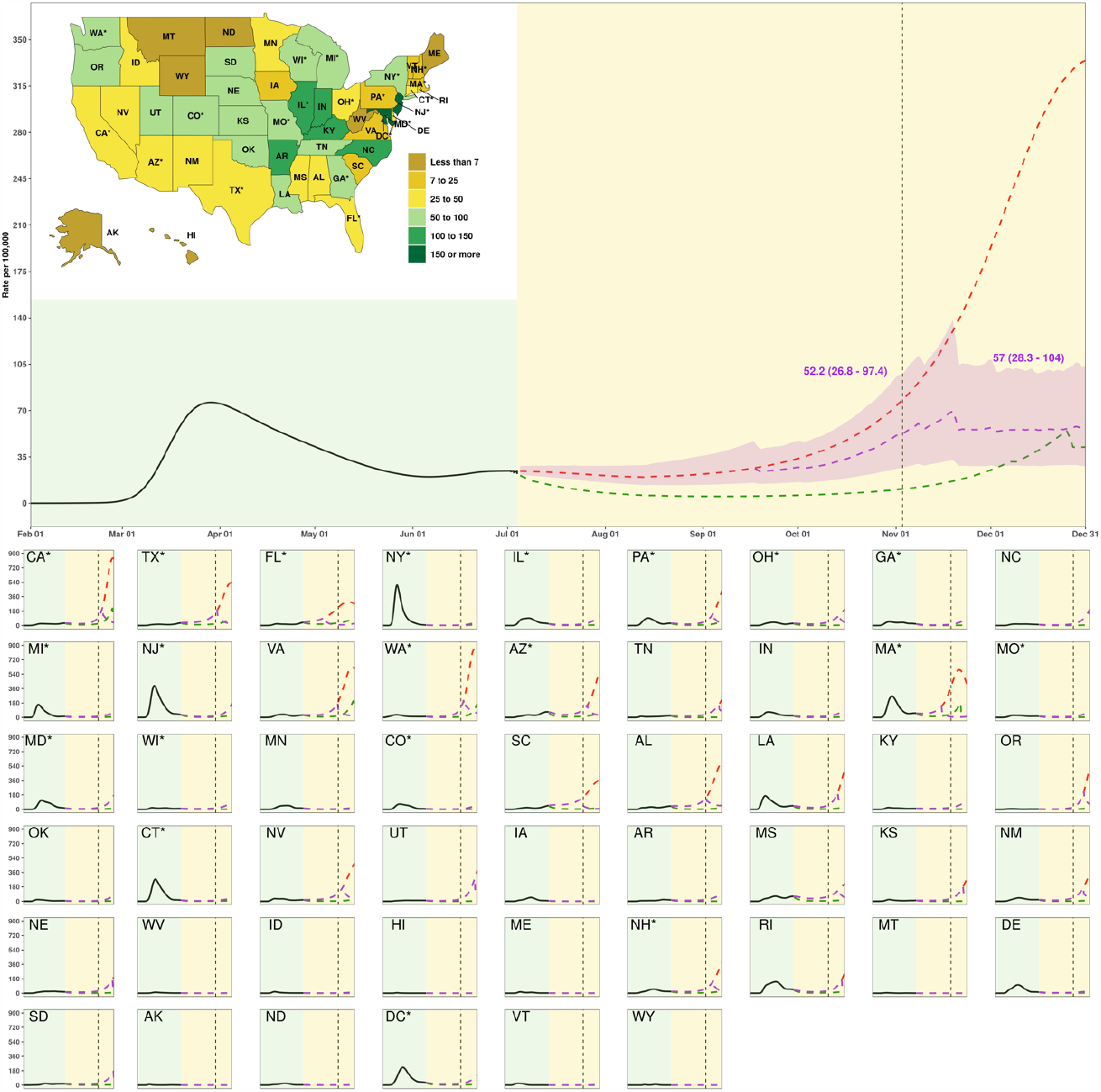
Estimated daily SARS-CoV-2 infection rate (per 100,000 population) by state for three scenarios. The inset map displays the estimated peak in daily infections from COVID-19 per 100,000 population by state between 04 July and 31 December. The light yellow background separates the observed and predicted part of the time series, before and after 04 July. The dashed vertical line identifies 03 November 2020. The red is the “mandates easing” scenario, the purple line the “plausible reference” scenario, and green line the “universal mask” scenario. Numbers are the means and uncertainty interval (UI) for the plausible reference scenario on dates highlighted. State panels are ordered by decreasing population size. Two-letter state abbreviations are provided in panels and the inset map. An asterisk next to state abbreviation indicates a state with one or more urban agglomerations exceeding two million persons. State panels are scaled to accommodate the state with the highest value (WA here), ranging from zero to 900. This map was generated with RStudio (R Version 3.6.3).

**EDF 9.**
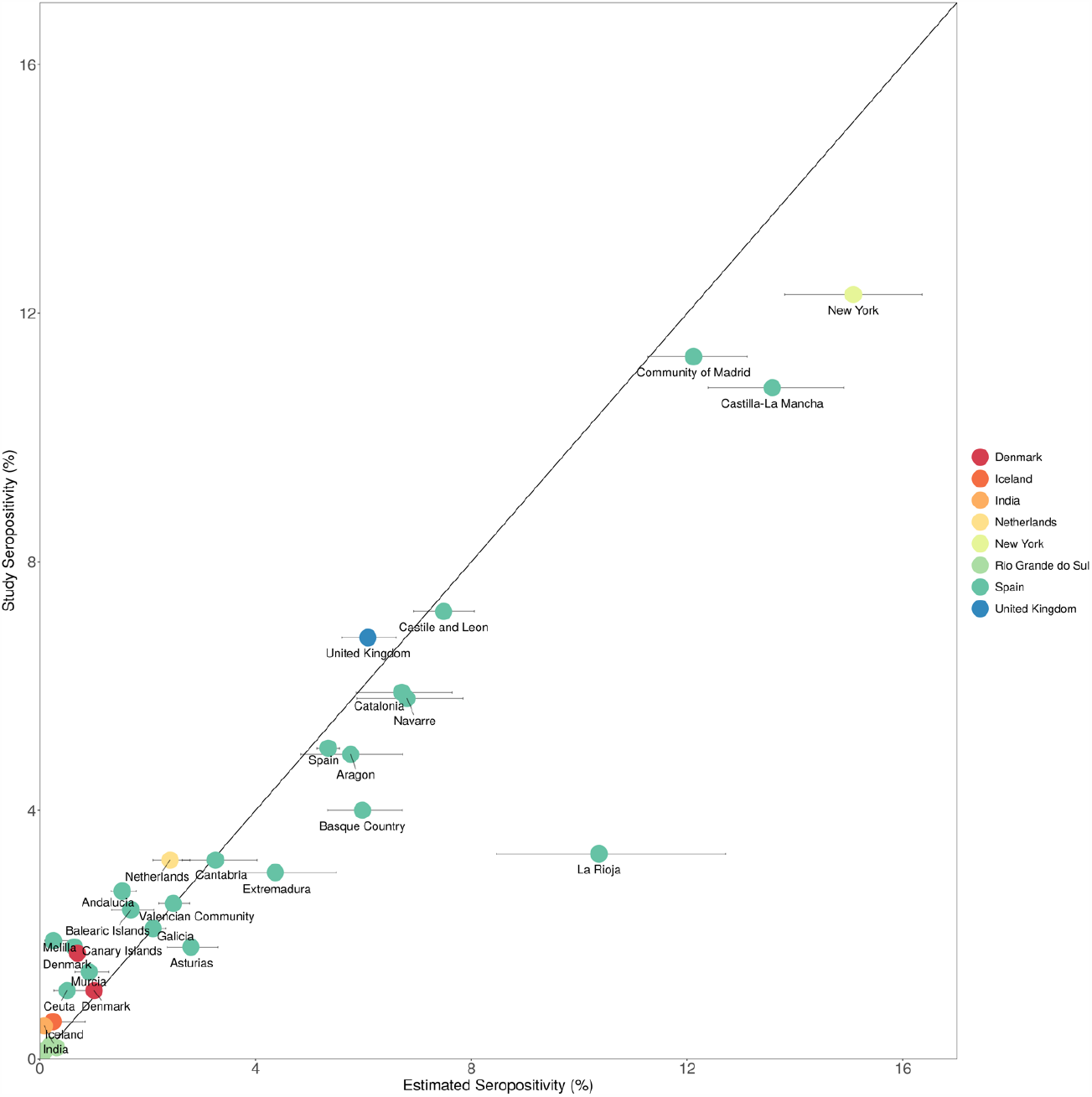
Modeled SARS-CoV-2 infection prediction totals compared with survey-derived seroprevalence rates in select locations.

